# Pre-Infection Mental Health, but Not Brain Volumetry, Predicts Risk of Post-COVID Condition: A Population-Based Cohort Study in the German National Cohort (NAKO)

**DOI:** 10.64898/2026.09.09.26362611

**Authors:** Claas Flint, Cord Spreckelsen, Christian Otto, Rafael Mikolajczyk, Jonas Frost, Luise Victoria Claaß, Jonathan Mathias Fasshauer, Markus Scholz, Laura Buschmann, Nicole Rübsamen, Marvin N. Wright, Lukas Burk, Fabian Bamberg, Christopher L. Schlett, Thoralf Niendorf, Michael Forsting, Yan Li, Karen Steindorf, Stefan Karrasch, Michael Leitzmann, Stefan Haug, Till W. Bärnighausen, Börge Schmidt, Manuela Harries, Vanessa Melhorn, Lilian Krist, Thomas Keil, Annette Peters, Nils Opel

## Abstract

**Background:** Post-COVID Condition (PCC) affected 10–30% of individuals after SARS-CoV-2 infection, and pre-infection predictors of who goes on to develop it are not well established. We compared pre-infection structural brain variation and baseline psychiatric phenotype as candidate predictors of PCC within a common multi-modal framework in the German National Cohort (NAKO). Both are candidate “first hits” under the second-hit hypothesis, which motivates the comparison; this is a prediction study, not a test of that hypothesis.

**Methods:** In 8,464 SARS-CoV-2-infected NAKO neuroimaging participants, of whom 2,304 (27.2%) met PCC criteria (weighted post-COVID syndrome (PCS) score > 10.75) and the rest were symptom-free controls, ten pre-infection modalities (T1-weighted volumetric brain MRI across six parcellations, baseline mental health (PHQ-9, GAD-7), demographics, and seven further biomedical and socioeconomic domains) entered a stacked ensemble with 10 × 5 nested cross-validation (primary metric: the area under the precision–recall curve, PR-AUC). Pre-specified analyses assessed transportability to a non-imaging cohort and robustness to the mental-health symptom trajectory and to unmeasured confounding. SARS-CoV-2 infection status and the PCC outcome were ascertained by self-report; no clinically confirmed diagnoses were available.

**Results:** Baseline mental health, assessed 3–8 years before infection, was the strongest predictor (38.0% of feature importance; standalone ROC-AUC = 0.640), whereas all six brain-MRI parcellations performed at or near chance (ROC-AUC 0.496–0.516). The multi-modal model achieved moderate, well-calibrated discrimination (ROC-AUC = 0.665, 95% CI 0.653–0.677; PR-AUC = 0.413, 0.392–0.434; ECE = 0.013). A four-modality non-imaging version, applied without retraining to NAKO participants without MRI, retained discrimination (ROC-AUC = 0.660, 0.654–0.665), meeting all three pre-specified equivalence criteria, the calibration intercept narrowly. The baseline mental-health odds ratio was 2.06 (1.83–2.32; E-value 2.66) and essentially unchanged under an alternative control definition (2.07); adjusting for the full mental-health symptom trajectory reduced it to a conservative lower bound of 1.38 (1.20–1.58) that remained independently significant. Baseline mental health did not predict objectively measured hyposmia in participants screened before their infection (0.89) while predicting self-reported smell loss in the same participants (1.92).

**Conclusions:** Baseline mental health years before infection is the strongest pre-infection predictor of PCC among the infected, and the association held across the pre-specified sensitivity analyses. Whether it acts specifically on COVID-19 sequelae is a separate question this design cannot answer, and the indirect evidence points away from specificity: the association is undiminished after mild infection but absent among the hospitalized, and baseline mental health predicts current mental-health symptom load slightly less strongly in infected than in non-infected participants. The volumetric structural candidate is not supported: prepandemic T1-weighted volumetry carried no predictive signal in this single neuroimaging cohort. Risk stratification may benefit from incorporating baseline psychiatric phenotype; whether treating it reduces PCC incidence requires interventional study.

## 1 Introduction

### 1.1 Post-COVID condition

Post-COVID Condition (PCC), also referred to as Long COVID, is defined by the World Health Organization (WHO) as a condition occurring in individuals with a history of probable or confirmed Severe Acute Respiratory Syndrome Coronavirus 2 (SARS-CoV-2) infection, usually three months from onset. The symptoms are new or persistent, last for at least two months, and cannot be explained by an alternative diagnosis (World Health Organization, 2021). Between 10% and 30% of infected individuals were affected during the pandemic, depending on population, outcome definition, and follow-up duration (Davis et al., 2023; Subramanian et al., 2022); a controlled meta-analysis of 50 studies and over 14 million participants confirms elevated risk versus uninfected comparators across 39 of 40 assessed symptoms (O’Mahoney et al., 2025). Symptoms range from fatigue and cognitive impairment to cardiorespiratory, musculoskeletal, and sensory complaints (Davis et al., 2023). Female sex is the most consistent risk factor; reports on age diverge, with some studies identifying younger age (Subramanian et al., 2022) and others older age (Bai et al., 2022) as risk-increasing.

Neurological and cognitive symptoms are among the most common and disabling PCC manifestations. Fatigue affects roughly 32% and cognitive impairment roughly 22% of those with post-acute sequelae, persisting beyond 12 weeks (Ceban et al., 2022). Elevated rates of neurological and psychiatric diagnoses (cognitive deficits, anxiety, mood disorders) have been reported in the six months after infection (Taquet et al., 2021). Even mild Coronavirus Disease 2019 (COVID-19) leaves objective deficits in memory and executive function months later (Hampshire et al., 2024). Whether pre-existing brain characteristics modulate this vulnerability remains an open question.

### 1.2 Neurobiological basis and the second-hit hypothesis

Converging evidence from neuroimaging, neuropathology, and molecular studies supports a neurobiological basis for PCC. SARS-CoV-2 can perturb Central Nervous System (CNS) homeostasis through direct neurotropism and, more prominently, indirect neuroinflammatory pathways (Meinhardt et al., 2021; Spudich & Nath, 2022), triggering neural cell dysregulation even after mild infection (Fernández-Castañeda et al., 2022). Longitudinal neuroimaging has demonstrated infection-associated reductions in gray matter thickness and increased tissue damage markers (Douaud et al., 2022), while large-scale epidemiological data document elevated long-term neurological risk (Xu et al., 2022).

On their own, these observations show only that infection can injure the CNS, which is equally consistent with a single-hit account in which the acute insult alone determines risk. What such an account does not explain is the pronounced inter-individual heterogeneity of PCC: only a minority of infected individuals develop persistent symptoms, and risk varies systematically with host factors such as sex (Bai et al., 2022; Davis et al., 2023; Subramanian et al., 2022). This heterogeneity motivates a *second-hit hypothesis* for PCC, adapting a framework originally proposed for neuropsychiatric vulnerability (Bayer et al., 1999) and drawing on recent mechanistic insights into COVID-19 neurobiology (Monje & Iwasaki, 2022). Pre-existing vulnerability constitutes a “first hit” that, when compounded by the neurotropic and neuroinflammatory insult of acute SARS-CoV-2 infection (the “second hit”), results in elevated risk for persistent neurological and cognitive symptoms. Observing that such a first hit predicts PCC among the infected is a necessary but not sufficient condition for the hypothesis, which additionally posits an interaction between vulnerability and infection that a single infected-only cohort cannot fully isolate.

Two concrete operationalizations of the first-hit dimension are plausible in a population cohort with pre-pandemic phenotyping. The *psychiatric* version posits that pre-existing depression, anxiety, and related affective disturbance constitute the relevant vulnerability and are captured by baseline psychometric assessment. The *structural* version posits that subclinical neuroanatomical variation, reduced neural reserve, or latent neuroinflammatory predisposition precede infection and are captured by pre-infection volumetric brain Magnetic Resonance Imaging (MRI). The two are not mutually exclusive: they can coexist, mediate each other, or both be wrong.

The psychiatric variant already has direct empirical support. A prospective three-cohort analysis by Wang et al. (2022) found that pre-infection depression, anxiety, worry, perceived stress, and loneliness were each associated with roughly 30–50% elevated PCC risk. Garjani et al. (2022) reported a similar pattern for pre-existing anxiety and depression in an early prospective specialty cohort. The structural variant has not been tested directly because the requisite combination of pre-pandemic neuroimaging and post-infection symptom follow-up has been rare. Elevated rates of neurological *and* psychiatric diagnoses after SARS-CoV-2 infection (Taquet et al., 2021), and the high prevalence of fatigue and cognitive impairment in post-acute sequelae (Ceban et al., 2022), are compatible with either version. Large population cohorts with both pre-infection imaging and pre-infection psychometrics allow a parallel, pre-specified test of the two operationalizations.

### 1.3 Predictive modeling of post-COVID condition

Machine Learning (ML) methods have been applied to predict PCC from clinical and demographic data, though the literature is heterogeneous with respect to outcome definitions, predictor sets, and analytical rigor. With *post-infection* predictors (acute-phase symptoms, hospitalization data, early healthcare utilization), models reach ROC-AUC (area under the receiver-operating-characteristic curve) values of 0.75–0.92 (Antony et al., 2023; Bergquist et al., 2024; Butzin-Dozier et al., 2024; Pfaff et al., 2022; Sudre et al., 2021), but such predictors are unavailable before infection and thus of limited use for prospective prevention. Acute-phase biomarkers such as viral load, Epstein-Barr virus reactivation, and specific autoantibodies similarly predict post-acute sequelae (Su et al., 2022).

*Pre-infection* models, the more relevant scenario for prospective risk stratification, report lower discrimination (ROC-AUC 0.60–0.66; Doni Jayavelu et al., 2026; Jin et al., 2023; Zang et al., 2024), though comprehensive electronic health record data can push values higher (Lee et al., 2025). These models rely mainly on demographics, comorbidities, and healthcare utilization.

Prior work has linked pre-infection psychiatric phenotype to elevated PCC risk in prospective, online-cohort, and claims-based designs (Bobak et al., 2024; Durstenfeld et al., 2023; Garjani et al., 2022; Greißel et al., 2024; Wang et al., 2022); however, to our knowledge no study has integrated pre-infection structural brain MRI and pre-infection psychometric assessment into a single multi-modal PCC prediction framework (Ahmad et al., 2024). A joint comparison of both candidate dimensions requires a cohort that carries both pre-pandemic volumetric MRI and validated psychometric assessment, as in the population cohort used here.

### 1.4 Study objectives

We compare structural brain MRI and baseline mental health as candidate pre-infection predictors within a common multi-modal predictive framework. The second-hit hypothesis motivates that comparison. We use data from the German National Cohort (NAKO), Germany’s largest population-based cohort study (German National Cohort, 2014), focusing on the neuroimaging subsample (see Section 2.1 for details).

Ten complementary pre-infection modalities enter a stacked ensemble: structural brain MRI across six atlas-based parcellations, baseline mental health, demographics, Socioeconomic Status (SES), cognitive function, physical activity, medical history, laboratory biomarkers, cardiovascular measures, and lung function. Structural brain MRI and baseline mental health are specified a priori as the two primary candidates. Either is considered unsupported as a pre-infection predictor if the corresponding modality fails to contribute predictive signal over and above demographic and socioeconomic baselines. Throughout, both SARS-CoV-2 infection status and the PCC outcome rest on participant self-report rather than clinically confirmed diagnoses, a constraint we return to in the Discussion.

## 2 Data

### 2.1 Study population and ethics

Data were obtained from the NAKO, a prospective population-based study designed to recruit 200,000 adults aged 20–69 years (German National Cohort, 2014); between 2014 and 2019 a total of 205,415 participants (ages 19–74 years) were examined at 18 study centers across Germany (Peters et al., 2022). The NAKO employs a two-level design for its baseline assessment (2014–2019): Level 1 was conducted in all participants, while Level 2, an extended examination program, was performed in a 20% subsample across all study centers. Structural brain MRI was offered at five of them; participants from six further centers were referred to these imaging sites, so the imaged participants come from eleven study centers in total.

The NAKO conducted two pandemic-related follow-up questionnaires: Corona-1 (2020) assessed initial infection status and the psychosocial impact of the first pandemic wave (Peters et al., 2020), and Corona-2 (sent in autumn 2022, answered until early 2023) collected detailed infection history, vaccination status, and current health status measures from all participants with a valid email address on record (Diexer et al., 2025; Mikolajczyk et al., 2024). Participants who reported a prior SARS-CoV-2 infection were additionally routed into a symptom section targeting the 4–12 month window after first infection: a filter question on persistent complaints, followed for those who affirmed it by 21 items covering fatigue, cognitive, cardiorespiratory, and other PCC-related domains.

Because the present study compares pre-pandemic brain MRI and baseline mental health as candidate preinfection predictors, the analytic sample was restricted to participants in the Level 2 neuroimaging subsample who also completed the Corona-2 follow-up. Participants were eligible for inclusion if they had structural brain MRI data acquired at baseline, completed the Corona-2 symptom assessment, had complete baseline demographic data (age, sex, study center), and had an observable symptom outcome. An outcome counts as observable when the symptom routing question^1^ was answered with either “yes” or “no”. Of the 117,461 Corona-2 completers, 19,242 also had structural brain MRI data. Of these, 7,917 did not report a SARS-CoV-2 infection. Among the 11,325 infected, the symptom outcome was unobservable for 2,365. For 1,846 the first infection was too recent for the 4–12 month reference window to have elapsed by the time of the survey, so the questionnaire routing did not administer the symptom section. All but three of them dated their first infection to August 2022 or later. A further 88 did not answer the routing question, and 431 answered fewer than 80% of the symptom items validly. This leaves 8,960 infected participants with brain MRI and an observable symptom outcome. The control arm is restricted to participants who explicitly declared being free of persistent post-infection symptoms at the Corona-2 routing question. Infected participants who endorsed post-infection symptoms but whose weighted Post-COVID Syndrome (PCS) remained at or below the moderate-severity threshold of 10.75 (*n* = 496 sub-threshold cases) therefore fall outside both arms of the primary analysis (clean-controls design; rationale in Section 2.2). The resulting analytic sample comprises *N* = 8,464 participants, of whom 2,304 (27.2%) meet PCC criteria (weighted PCS > 10.75; Bahmer et al., 2022). Figure 1 summarizes the participant flow.

**Figure 1:**
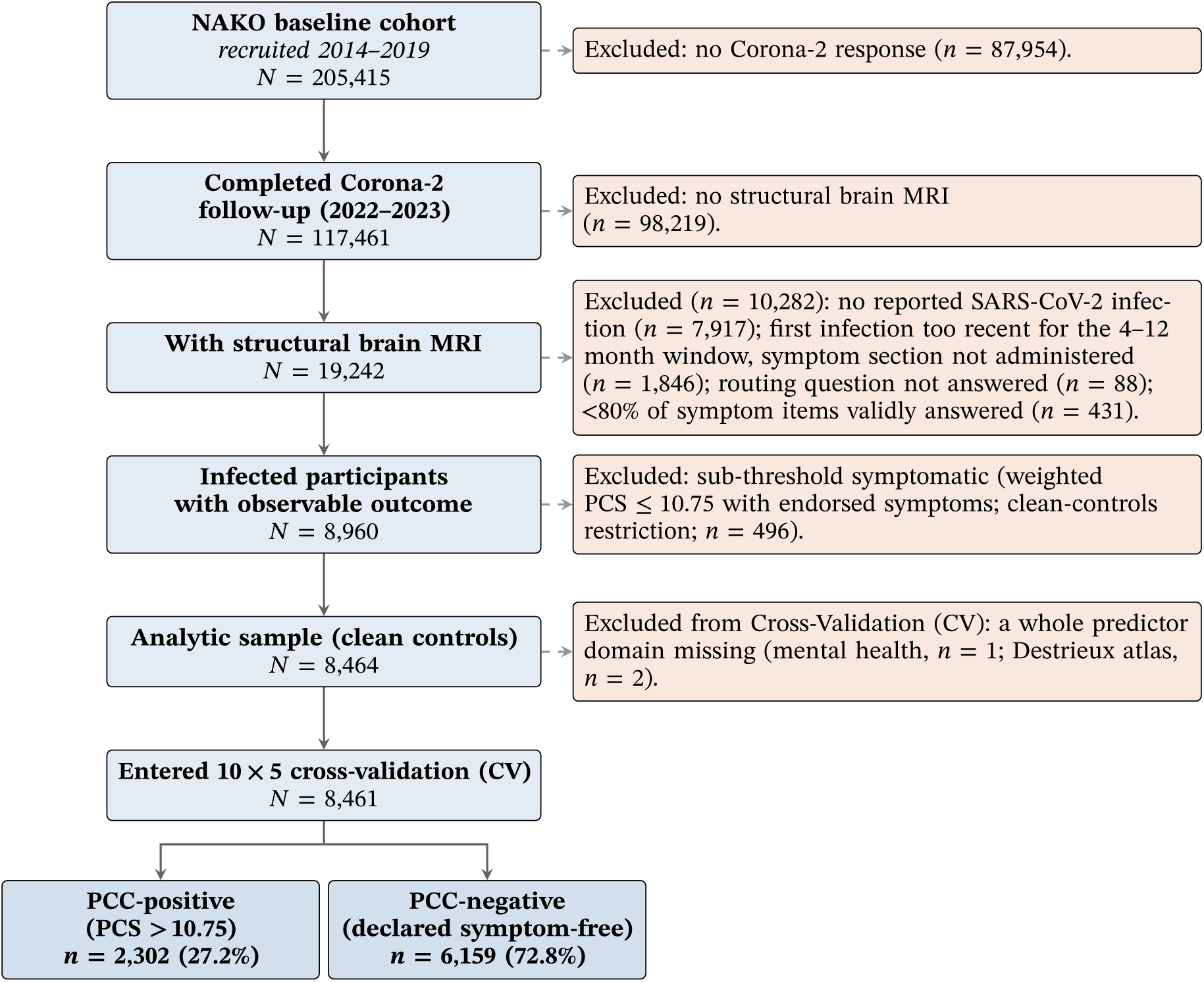
Participant flow. Participant flow from the NAKO baseline cohort to the analytic sample. Boxes on the main axis (blue) show included participants at each stage; boxes to the right (orange, dashed) summarize exclusions. The analytic sample uses a clean-controls definition that excludes sub-threshold symptomatic participants from both arms (see Section 2.2). Three participants with a whole modality missing (one mental health, two Destrieux atlas) are excluded from cross-validation, which therefore runs on *N* = 8,461 rather than the analytic-sample *N* = 8,464. NAKO = German National Cohort; MRI = Magnetic Resonance Imaging; SARS-CoV-2 = Severe Acute Respiratory Syndrome Coronavirus 2; PCC = Post-COVID Condition; PCS = Post-COVID Syndrome.

The NAKO is performed with the approval of the relevant local ethics committees and is in accordance with national law and with the Declaration of Helsinki of 1975 (in the current, revised version). Written informed consent was obtained from all participants. Data collection and storage comply with the European Union (EU) General Data Protection Regulation (GDPR). All analyses were conducted on pseudonymized data with restricted access following NAKO data access policies.

### 2.2 Outcome definition

PCC was assessed using 21 symptom items derived from the WHO clinical case definition (World Health Organization, 2021) and established PCC symptom inventories (Diexer et al., 2025). Participants who reported a prior SARS-CoV-2 infection were first asked a single routing question, namely whether they had experienced at least one persistent complaint in the *4–12 month window after their first infection*; only those answering “yes” were then shown the 21 individual items. Anchoring both stages to that window means symptom endorsement reflects infection-related sequelae rather than concurrent illness or pre-existing conditions. The items covered fatigue and exercise tolerance (fatigue or exhaustion, reduced physical capacity), cognition (memory or word-finding problems, concentration difficulties), cardiorespiratory symptoms (shortness of breath, chest tightness or pain, cough, cardiac complaints such as palpitations, circulatory complaints such as dizziness on standing), neurological symptoms (headache; tingling, burning or numbness), smell and taste disturbance, sleep problems, joint or muscle pain, and other symptoms (fever, sweating, runny or blocked nose, gastrointestinal complaints, loss of appetite or weight, hair loss). Each item was scored as binary (0 = absent, 1 = present).

The primary PCC outcome was defined as the weighted post-COVID syndrome (PCS) score proposed by Bahmer et al. (2022). The score is a 0–59 point aggregate of 12 empirically derived symptom complexes weighted by their epidemiological association with persistent post-infection morbidity (fatigue 7.0, cough/wheeze 7.0, neurological 6.5, joint/muscle pain 6.5, ear, nose, and throat (ENT) 5.5, gastrointestinal 5.0, sleep disturbance 5.0, exercise intolerance 4.0, infection signs 3.5, chemosensory 3.5, chest pain 3.5, dermatological 2.0). Each complex contributes its weight if any constituent symptom was endorsed; the total is summed across complexes. We assigned the NAKO items to the complexes by content. Two of the 21 items, cardiac complaints and circulatory complaints, have no counterpart among the twelve complexes and do not enter the score; they still count toward the symptom-count definition below. A participant was classified as PCC-positive when the weighted PCS exceeded 10.75, corresponding to the “moderate-or-worse” severity cut-off reported by Bahmer et al. (2022).

The weighted PCS is built from the same 21-item inventory as the symptom-count definition used in related German COVID-19 cohort analyses (Diexer et al., 2025), but is the stricter of the two criteria. Among the 8,960 infected participants with an observable outcome, 2,764 endorsed at least one symptom whereas 2,304 exceeded the weighted cut-off. The 496 participants who separate the two definitions are exactly the sub-threshold group that the clean-controls restriction described below removes from both arms. Of them, 460 endorsed at least one symptom without reaching the cut-off, and 36 answered the routing question with “yes” but then endorsed none of the 21 items. Within the analytic sample the two definitions therefore label the same participants, because every retained control declared no post-infection symptoms at the routing question.

Within the infected analytic sample we further restrict the control arm to participants who answered the routing question with “no”. Those who answered “yes” are retained only if their weighted PCS exceeds the moderate-severity cut-off (> 10.75, PCC-positive); the 496 sub-threshold participants are excluded. The rationale is that a binarized cut-off on a continuous severity score creates a gray zone of mildly symptomatic participants whose biological state is neither a clean case nor a clean control. Assigning them to the control arm makes the control distribution bimodal and partially overlaps the case distribution on the very predictors we want to measure, systematically attenuating effect-size estimates in a hypothesis-testing framework (Altman & Royston, 2006; Steyerberg, 2019). With a sample size of several thousand per target, the power loss from excluding the sub-threshold group is negligible relative to the bias it would introduce under a mixed-controls definition. A sensitivity analysis treating the sub-threshold group as PCC-negative (*N* = 8,960, prevalence 25.7%) left the baseline mental-health odds ratio essentially unchanged (2.07 [1.84–2.33]; Section 5.2).

### 2.3 Predictor variables and missing data

Ten complementary data modalities collected at NAKO baseline were integrated as predictor variables (Table 1). The predictor domains were fixed before the analysis on three rules. Brain MRI and baseline mental health were set a priori, because they are the two candidate pre-infection dimensions this study exists to compare. The remaining eight cover the domains the NAKO baseline examination program measures in the full cohort, so that the two candidate predictors are judged against the breadth of what a population cohort routinely records rather than against a hand-picked comparison set. Variables measured after the infection, and variables whose content overlaps the symptom inventory that defines the outcome, were excluded throughout to avoid circularity; this is why the Corona-2 psychometric scores appear in the sample description but never as predictors. The available breadth was additionally bounded by the approved data export (Application No. NAKO-882). Central to the study design, structural brain MRI features comprise regional volumes from six complementary atlases, each treated as a separate sub-modality to capture distinct neuroanatomical organizing principles. The remaining nine modalities span demographics, SES, cognitive function, physical activity, medical history, laboratory values, cardiovascular measures, lung function, and mental health; feature-level composition and counts are summarized in Table 1.

**Table 1:** Predictor modalities. Summary of predictor modalities within the neuroimaging subsample (*N* = 19,242; the full Level-2 neuroimaging subsample, before the infection/outcome and clean-controls restrictions that define the analytic sample of *N* = 8,464 and the cross-validation sample of *N* = 8,461). All variables were assessed at NAKO baseline (2014–2019) and have participant-level availability, though individual modalities exhibit varying rates of systematic missingness (see Supplementary Table S2). Feature counts for non-MRI modalities reflect variables after domain-specific preprocessing, including removal of redundant features (e.g., raw variables subsumed by derived measures); counts for demographics and SES refer to the number of input variables before one-hot encoding of categorical features (see Supplementary Table S2 for raw feature counts). Brain MRI features were extracted using FreeSurfer 7.1 from T1-weighted 3 T scans; feature counts reflect raw variables entering the pipeline before PCA dimensionality reduction.

| Modality | Features | Description |
| --- | --- | --- |
| *Brain MRI sub-modalities* |  |  |
| Desikan-Killiany | 68 | Regional volumes (cortical parcellation, 34 regions × 2 hemispheres) |
| Destrieux | 187 | Regional volumes (detailed cortical parcellation, gyri and sulci) |
| Julich | 50 | Regional volumes (cytoarchitectonic areas) |
| Subcortical | 186 | White matter and subcortical structure volumes |
| Yeo Networks | 96 | Functional network volumes (7-network parcellation) |
| Cerebellar | 12 | Cerebellar lobule volumes |
| Demographics | 3 | Age, sex, study center |
| SES | 27 | ISCED, ISEI prestige, employment, income, household size |
| Cognitive | 8 | Word list recall, verbal fluency, Stroop, digit span, Purdue pegboard |
| Physical activity | 20 | MET-minutes/week (QUAP and GPAQ), occupational activity, sedentary time, active transport |
| Medical history | 35 | Anthropometry, disease indicators including hypertension, medications, infections, cardiovascular procedures, smoking |
| Laboratory | 19 | Lipids, glucose and HbA1c, CRP, liver/kidney/thyroid function, electrolytes, blood count |
| Cardiovascular | 7 | Blood pressure, heart rate, PWV, augmentation index, ABI |
| Lung function | 4 | FEV1%pred, FVC%pred, PEF, FEF<sub>25–75</sub> |
| Mental health | 21 | PHQ-9 sum and nine items, GAD-7, MINI depression, panic, stress, onset/duration |

MRI = Magnetic Resonance Imaging; SES = Socioeconomic Status; ISEI = International Socio-Economic Index; MET = Metabolic Equivalent of Task; QUAP = NAKO physical activity questionnaire; GPAQ = Global Physical Activity Questionnaire; HbA1c = Glycated Hemoglobin; CRP = C-Reactive Protein; PWV = Pulse Wave Velocity; ABI = Ankle-Brachial Index; FEV1 = Forced Expiratory Volume in 1 second; FVC = Forced Vital Capacity; PEF = Peak Expiratory Flow; FEF<sub>25–75</sub> = forced expiratory flow at 25–75% of FVC; PCA = Principal Component Analysis; ISCED = International Standard Classification of Education; PHQ-9 = Patient Health Questionnaire-9; GAD-7 = Generalized Anxiety Disorder 7-item scale; MINI = Mini-International Neuropsychiatric Interview.

Structural MRI was acquired in the Level 2 subsample at five imaging study centers using 3 T Siemens Skyra scanners (Siemens Healthineers, Erlangen, Germany) according to a standard operating procedure (Bamberg et al., 2015, 2024) (*n* = 19,242); FreeSurfer 7.1 extracted regional brain volumes under six complementary parcellation schemes (Table 1). Trained personnel administered cognitive assessments under standardized protocols. Physical activity was quantified via two parallel self-administered instruments, both covering the preceding 12 months: the NAKO physical activity questionnaire (QUAP), which contributes the activity-domain detail (occupational activity, active transport, sport, sedentary time), and the Global Physical Activity Questionnaire (GPAQ), from which we take the total MET-minutes/week. The two are carried as separate features rather than pooled, because they agree only moderately (Spearman *ρ* = 0.38 among participants who completed both) and differ substantially in coverage. Blood was drawn after overnight fasting and analyzed at central laboratories; blood pressure was recorded with automated oscillometric devices. Spirometry followed American Thoracic Society (ATS)/European Respiratory Society (ERS) guidelines. Smoking entered the medical-history modality as NAKO’s derived tobacco variables: smoking status (never, former, current), pack-years, years smoked, and the lifetime average number of cigarettes per day. NAKO derives the last only for current smokers; we set it to zero for never smokers and left it missing for former smokers, whose values the imputation fills. Age at cessation is also extracted but is defined only for former smokers, so at 69% missing it falls to the missingness filter described in the Methods and does not reach the classifier. Baseline mental health was assessed via the PHQ-9, GAD-7, and the Mini-International Neuropsychiatric Interview (MINI) depression module; the nine PHQ-9 items enter alongside the sum score. PHQ-9 and GAD-7 sum scores are complete-case, following NAKO’s own derivation, which applies no prorating and tolerates no missing item (Streit et al., 2023).

Nearly every participant in the analytic sample has data in all ten predictor modalities; a whole modality is missing for at most 7.1% of participants (lung function), and features vary in missingness rates (Table 1; Supplementary Table S2). Sporadic missing values were handled by modality-specific imputation (see Methods). Restricting to the neuroimaging subsample introduces minor selection effects (younger, less often female, slightly lower depressive symptom burden), while PCC prevalence is indistinguishable between Corona-2 completers with and without MRI (Supplementary Table S3).

## 3 Methods

### 3.1 Analytical framework

We developed a multi-modal meta-learning framework to predict PCC from the ten baseline data modalities described in Section 2.3 (Table 1). The stacking architecture addresses three problems: controlling for demographic confounding without losing predictive signal, handling heterogeneous data types with varying dimensionality and missingness patterns, and preventing overfitting through nested cross-validation and regularized base learners.

The pipeline follows a two-stage architecture (Figure 2). In the first stage, each modality was processed through a modality-specific pipeline that includes preprocessing and training of a base learner. Demographics (age, sex, study center) occupy a special role: rather than being orthogonalized themselves, they are the confounders against which all other modalities were adjusted via the first stage of Double Machine Learning (DML) (Chernozhukov et al., 2018). Specifically, we applied X-residualization only. Cross-fitted eXtreme Gradient Boosting (XGBoost) nuisance models (5-fold CV, refit within each outer training fold to prevent leakage across the outer cross-validation boundary) partial out demographic associations from each feature. The outcome *y* was not residualized, because the demographics base learner directly captures confounder-to-outcome effects within the stacking architecture. This captures nonlinear demographic effects and interactions without manual specification, removes demographic associations from all features while preserving outcome-relevant signal, and avoids orthogonalizing demographics against themselves.

**Figure 2:**
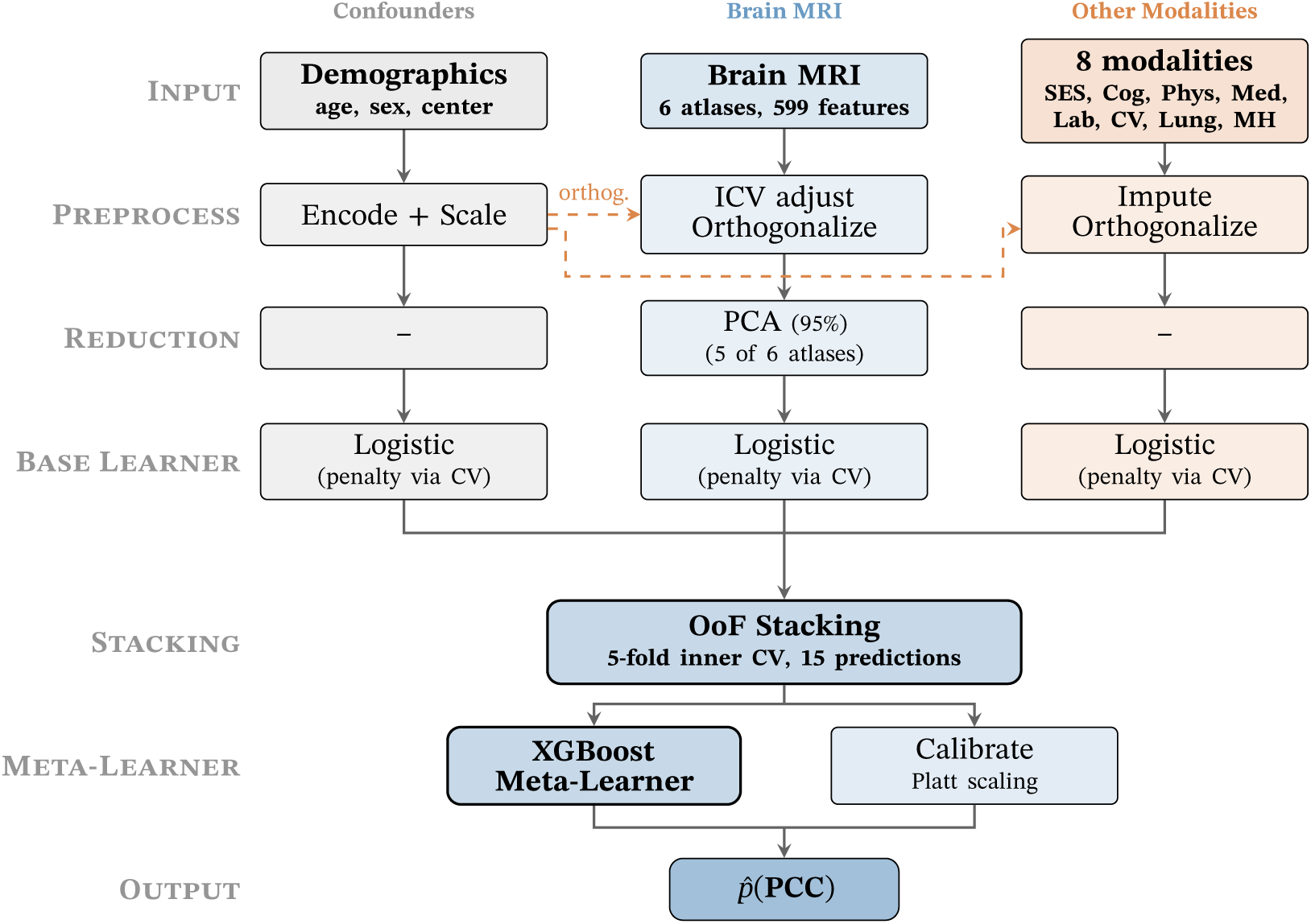
Multi-modal meta-learning pipeline. Three modality groups are processed through parallel vertical pipelines. **Demographics** (gray) are the confounders and follow a direct path without orthogonalization. **Brain MRI** features (blue) from six atlases undergo ICV adjustment via the residual method, orthogonalization against demographics (dashed arrows), PCA (for atlases with > 20 features), and penalized logistic regression. **Other modalities** follow analogous preprocessing without a reduction step and logistic regression base learners. All base learners select the penalty type (from a per-modality set of L2, L1 and elastic net) and regularization strength via 3-fold inner CV. OoF predictions from 5-fold inner CV (15 total: 6 MRI atlases + 1 demographics + 8 other) are combined via an XGBoost meta-learner with Platt calibration. MRI = Magnetic Resonance Imaging; ICV = Intracranial Volume; PCA = Principal Component Analysis; CV = Cross-Validation; OoF = Out-of-Fold; SES = Socioeconomic Status.

Each non-demographic modality underwent a four-stage pipeline: (1) domain-specific preprocessing, (2) *k*-nearest-neighbor imputation of sporadic missing values (*k* = 5), (3) orthogonalization against demographics via DML, and (4) z-score standardization. Preprocessing encoded categorical variables, removed redundant features such as raw variables subsumed by derived measures or hierarchical codes subsumed by summary indices, and filtered features exceeding 50% missing values. Values outside predefined plausibility ranges had already been set to missing when the data were extracted. The imputation neighborhood (*k* = 5) is the common default and was not tuned. For MRI volumes, the preprocessing stage additionally applied Intracranial Volume (ICV) adjustment via the residual method, where FreeSurfer’s estimated total intracranial volume entered the orthogonalization step as an additional covariate; this avoids the double sex correction inherent to the proportion method. MRI atlases followed a separate pipeline: five atlases with more than 20 features (Desikan-Killiany, Destrieux, Julich, subcortical, Yeo networks) underwent z-score standardization followed by Principal Component Analysis (PCA) (retaining components explaining 95% of variance) and re-standardization of retained components. Cerebellar volumes (12 features) were used directly. No modality passed through a separate feature-selection step; regularization in the base learner, with the penalty chosen by inner cross-validation, takes that role for all modalities.

Following preprocessing, each modality trained a dedicated penalized logistic-regression base learner. Penalty type (L2, L1, or elastic net) and regularization strength were selected via 3-fold inner CV optimizing average precision; all base learners used balanced class weights. Solver choice, scoring rationale, and per-modality *C* grids are reported in Supplementary Table S1.

### 3.2 Meta-learning and calibration

In the second stage, Out-of-Fold (OoF) predictions from all 15 base learners (ten modality groups, with six brain MRI atlases treated separately) were combined by a meta-learner. Traditional stacking (Wolpert, 1992) trains the meta-learner on in-sample predictions, which invites overfitting. We avoided this with nested OoF stacking (Varma & Simon, 2006): for each outer CV fold, base learners were trained on the training portion via inner CV and generated predictions on held-out inner validation sets. These predictions, concatenated across inner folds to cover the full outer training set, form the meta-learner’s training data and reflect realistic base learner performance (Figure 3).

**Figure 3:**
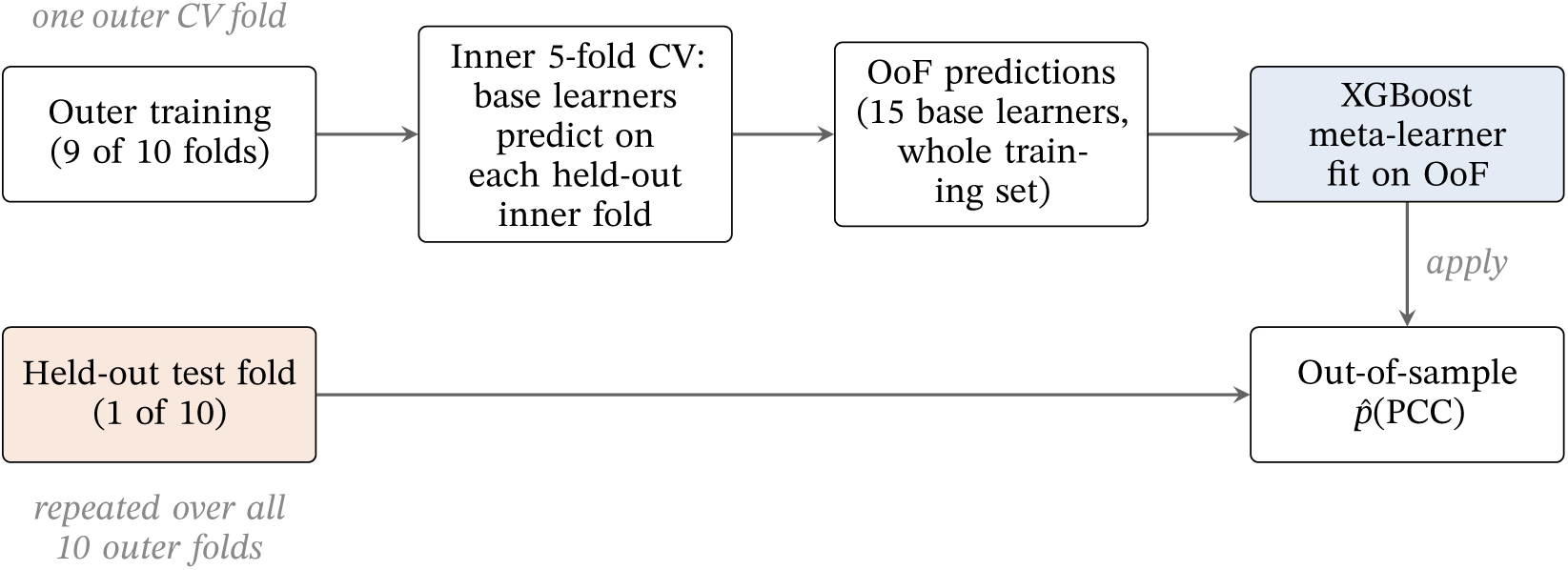
Nested out-of-fold stacking. For each of the 10 outer cross-validation folds, the nine outer-training folds pass through an inner 5-fold cross-validation in which every modality’s base learner is trained on four inner folds and predicts on the held-out fifth. Concatenating these held-out predictions yields OoF predictions that cover the entire outer-training set without any base learner ever scoring its own training data. The XGBoost meta-learner is fit on these OoF predictions and then applied once to the outer test fold, so the reported discrimination metrics are out-of-sample at both the base-learner and meta-learner stages. OoF = Out-of-Fold; CV = Cross-Validation; PCC = Post-COVID Condition.

We used XGBoost (Chen & Guestrin, 2016) as the meta-learner because its sparsity-aware split finding natively handles missing inputs, enabling the passthrough strategy for systematically missing modalities described below. Hyperparameters were tuned via randomized search (50 iterations) with internal 5-fold CV on the OoF training predictions, optimizing PR-AUC (area under the precision–recall curve; Supplementary Table S1). The meta-learner’s predictions were calibrated using Platt scaling (Platt, 1999) with 3-fold CV.

Binary PCC prevalence of 27.2% in the infected analytic sample creates class imbalance, which we addressed through balanced class weights in all classifiers and use of PR-AUC as the primary evaluation metric. Because a summary discrimination measure says nothing about how the model behaves at a decision point, sensitivity, specificity, Positive Predictive Value (PPV) and Negative Predictive Value (NPV) at two operating points are reported alongside it in Table 7. Missingness varied across modalities (Supplementary Table S2), and was addressed at two levels. Sporadic missingness (individual features absent within an otherwise observed modality) was handled via *k*-nearest-neighbor imputation (*k* = 5). Systematic missingness, where all features are absent for a modality (highest in lung function at 7.1%, followed by laboratory values, cognitive function, and the Desikan-Killiany atlas), was handled by passing NaN directly to the XGBoost meta-learner, avoiding fabrication of uninformative predictions.

### 3.3 Evaluation

Model performance was evaluated using PR-AUC as the primary metric, supplemented by ROC-AUC, Brier score (Brier, 1950), and, at the operating points, the F1 score (the harmonic mean of PPV and sensitivity). Calibration was assessed by slope, intercept (Van Calster et al., 2016), and Expected Calibration Error (ECE) computed with 10 equal-count (quantile) probability bins, and clinical utility by sensitivity at the 90% specificity threshold, PPV, NPV, and Decision Curve Analysis (DCA) (Vickers & Elkin, 2006) evaluated across threshold probabilities from 1% to 99%. Uncertainty for primary discrimination metrics was quantified via bootstrap (1,000 iterations) for 95% Confidence Intervals (CIs). Resampling was stratified by outcome for ROC-AUC, which is invariant to outcome prevalence, and unstratified for PR-AUC, which is not: holding the observed prevalence fixed would suppress a genuine source of variation and return an interval that is too narrow. Significance was assessed via a label-permutation test (1,000 iterations) on the fixed OoF predictions, recomputing PR-AUC against the unchanged predictions under shuffled labels (Supplementary Section S3).

Individual modality contributions were assessed through incremental performance (ΔPR-AUC when added to a demographics-only baseline), modality masking (ΔPR-AUC when replacing the modality’s predictions with NaN in the fitted model), and SHapley Additive exPlanations (SHAP) values (Lundberg & Lee, 2017) computed via TreeExplainer (Lundberg et al., 2020) on the uncalibrated XGBoost meta-learner. The incremental and SHAP analyses operate on the OoF training predictions and therefore provide meta-learner-level attribution; the masking analysis is computed per outer fold on held-out participants, as are the primary discrimination metrics (ROC-AUC, PR-AUC). Because the XGBoost meta-learner routes missing inputs to a learned default direction rather than to an imputed value (Chen & Guestrin, 2016), the NaN masking measures each modality’s contribution under the model’s learned handling of its absence rather than via random permutation. We therefore read it alongside the incremental and SHAP analyses rather than in isolation. Statistical comparisons against the mental-health baseline used Nadeau–Bengio corrected paired tests (Nadeau & Bengio, 2003) with Holm adjustment. Masking one atlas at a time understates what six correlated atlases contribute jointly, because the remaining five still carry the masked atlas’s information. We therefore also removed all six together within each outer fold, both by masking them with NaN under the fold’s own meta-learner and by retraining the fold’s meta-learner on the training-fold OoF predictions of the nine remaining modalities with the identical hyperparameter search and seed, and scored the held-out fold in both cases; retraining stops at the meta-learner because each base learner sees only its own modality. The criterion, fixed before this analysis was run but after the per-atlas MRI results were known, is equivalence: the 95% CI of the held-out difference (full model minus model without MRI) from a paired participant bootstrap (1,000 iterations, stratified for ROC-AUC and unstratified for PR-AUC) had to lie entirely within ±0.03 on both metrics, the margin of the transportability test (Section 3.4), with a Nadeau–Bengio test over the 10 per-fold differences as secondary criterion.

The outer cross-validation used *k* = 10 stratified folds and the inner loop, which produces the out-of-fold predictions and tunes the meta-learner, used *k* = 5 folds (base-learner penalties are tuned in a further 3-fold split); the 10 × 5 scheme provides df = 9 for Nadeau–Bengio corrected paired tests of modality-level differences while keeping the per-fold training size large enough for stable base-learner fitting.

As sensitivity analyses, we assessed robustness of the primary outcome along three axes: outcome operationalization, control-arm definition, and demographic orthogonalization. First, we compared the weighted PCS against a hypothesis-aligned neurocognitive subtype, defined as endorsement of at least two of four items spanning fatigue, reduced physical capacity, memory problems, and concentration problems. Second, we evaluated an alternative *mixed-controls* definition that retains sub-threshold symptomatic participants (symptom-endorsing with weighted PCS ≤ 10.75) as PCC-negatives, to verify that the clean-controls primary design does not trivially amplify effects. Third, we tested a non-orthogonalized pipeline (DML disabled) to determine whether the first-stage X-residualization masks age– or sex-mediated vulnerability effects that the orthogonalized primary design would remove. The neurocognitive-subtype threshold is ≥ 2 of four items, yielding a syndrome-like definition empirically distinct from the primary outcome (*κ* ≈ 0.786; Supplementary Table S5); a one-item threshold would, within the clean-controls design, coincide with the weighted PCS by construction and therefore provide no independent sensitivity signal. Restricting the outcome to domains most plausibly mediated by pre-existing cerebral vulnerability additionally tests whether the mental-health signal is sharpened when somatic complaints that the broader PCS aggregates are removed.

As a positive control on the imaging pipeline itself, we verified that the processed MRI features carry usable information independently of the PCC task. We predicted participants’ baseline age (RidgeCV regression) and sex (L2-penalized logistic regression) from the same atlas features, before orthogonalization, under 10-fold out-of-fold cross-validation on the identical analytic sample (Supplementary Table S10).

Six further robustness analyses, all using the composite baseline-MH-positive exposure adjusted for age, sex, and center and fitted outside the prediction pipeline, assess reporting style, infection specificity, effect modification and behavioral confounding. (i) *Chemosensory outcome:* we refit the logistic regression with a binary chemosensory outcome (post-infection loss of smell or taste), the one PCC item with an instrument counterpart in NAKO (see (vi)). (ii) *Difference-in-association:* on the full Corona-2 sample (infected and non-infected, who all completed the current PHQ-9/GAD-7) we tested a baseline-MH × infection interaction on current MH-positivity, as a coarse probe of infection-driven amplification. (iii) *Mixed-controls sensitivity:* we recomputed the baseline-MH odds ratio under the alternative outcome that retains sub-threshold symptomatic participants as PCC-negative. (iv) *Sex modification:* we assessed whether the baseline-MH effect differs by sex, fitting a multiplicative MH × sex interaction (tested by Wald and likelihood-ratio tests) and sex-stratified logistic models on the same composite exposure. (v) *Tobacco confounding:* we added smoking status (never as reference, former and current as indicators) and pack-years to the adjustment set. Smoking is associated with both depressive symptoms and general health and is therefore a candidate explanation rather than merely a further predictor. Tobacco data are incomplete for 1.0% of the analytic sample, almost all of them because NAKO records the smoking status as unknown; those participants were excluded rather than folded into the never-smoker reference, which would enlarge that group with people who may well have smoked. The reference model for the comparison is therefore the primary model refitted on the participants with complete tobacco data (*n* = 8,375), so that the difference is attributable to the adjustment rather than to the change of sample. (vi) *Objective olfactory anchor:* because the chemosensory item in (i) is the one PCC symptom NAKO also measures with an instrument, we set the two against each other in the same participants. The instrument is the 12-item Sniffin’ Sticks identification screening administered at the Level 2 follow-up examination, which NAKO reads into a normosmia, hyposmia, or anosmia classification; we use that classification rather than a threshold of our own, and treat as missing the screenings NAKO declines to classify. Participants reporting a cold in the six weeks before the test are excluded, since nasal congestion depresses the identification score for reasons unrelated to the exposure; an unrecorded cold counts as no cold, and the flag is entered as a covariate in a sensitivity panel instead. The examination year is reconstructed as the baseline visit year plus the baseline-to-examination age difference, which resolves the visit to the calendar year and is available for part of the cohort. That splits the infected clean-controls sample into those screened before their reported infection, for whom the instrument cannot register post-COVID-19 anosmia, and those screened after it, for whom it can. Participants screened in their year of infection cannot be ordered against it and enter neither stratum. Within each stratum we fit the objective and the self-reported outcome on identical participants with identical adjustment, and obtain a percentile interval for the ratio of the two odds ratios from 1,000 participant bootstrap resamples. A linear-probability model regressing the self-report on the instrument reading and the baseline PHQ-9 together, and the same objective model among the never-infected, complete the panel (Supplementary Table S15).

To bound the baseline-mental-health effect against unmeasured confounding, we computed an E-value (VanderWeele & Ding, 2017) from the age-, sex-, and center-adjusted odds ratio of a dichotomous MH-positive indicator (PHQ-9 sum ≥ 10, GAD-7 sum ≥ 10, or MINI-based major depression). The E-value was computed at both the point estimate and the lower 95% CI bound (Supplementary Section S4).

As a complementary construct-persistence sensitivity, we refit the same logistic regression with an additional Corona-2 current-MH indicator (PHQ-9 sum ≥ 10 or GAD-7 sum ≥ 10 computed from the PHQ and GAD items administered at the Corona-2 assessment) entered as a covariate. We report the odds ratio with and without this adjustment and the log-odds-coefficient shrinkage. To extend this beyond the two-wave comparison, we additionally fit a three-wave trajectory model. It enters PHQ/GAD-derived MH-positive indicators at all three available NAKO assessments simultaneously: baseline (T0, 2014–2019; with the MINI added because it is administered only at baseline), the Corona-1 mail-in questionnaire (T1, 2020), and Corona-2 (T2, 2022–2023). The Corona-1 indicator was reconstructed from the raw PHQ-9 and GAD-7 items of the Corona-1 questionnaire, rescaled from the NAKO 1–4 encoding to the canonical 0–3 scale. This trajectory model partitions the baseline-MH association into a part shared with intermediate (T1) and concurrent (T2) mental health and a residual that pertains only to the pre-pandemic phenotype.

To address the construct-overlap concern that PHQ-9/GAD-7 share somatic items (fatigue, sleep disturbance, concentration difficulty) with the Corona-2 symptom inventory, we decomposed the 21-item symptom set into two groups and refit the baseline-MH logistic regression on each as a secondary binary outcome. Five items with direct construct overlap with Diagnostic and Statistical Manual of Mental Disorders (DSM)-5 Major Depressive Disorder (MDD)/Generalized Anxiety Disorder (GAD) somatic criteria (fatigue, concentration problems, memory problems, sleep problems, loss of appetite) formed the *overlap* set. The remaining 16 items, aligned with COVID-19 pathophysiology (smell/taste loss, fever, respiratory symptoms, cardiovascular symptoms, gastrointestinal symptoms, nerve problems, hair loss, musculoskeletal pain, reduced physical capacity, sweating, headache, and circulation problems), formed the *mechanism-aligned* set. Each outcome was defined as the endorsement of at least one symptom in the corresponding set. We complemented the binary models with negative-binomial Generalized Linear Models (GLMs) on symptom counts to verify that the decomposition does not hinge on the ≥ 1-item threshold. If baseline-MH predicted PCC chiefly via symptom-construct overlap, the odds ratio for the overlap outcome should materially exceed that for the mechanism-aligned outcome.

### Mental-health modality composition

The primary multi-modal model treats baseline mental health as a single modality whose feature set spans the PHQ-9 sum score and its nine constituent items, GAD-7, MINI-based major-depression diagnostics, the PHQ-Panic items, and the PHQ-Stress items. To localize which instrument carries the dominant mental-health signal, we additionally fit the entire pipeline with the mental-health modality split into five instrument-specific sub-modalities (PHQ-9, GAD-7, MINI, PHQ-Panic, PHQ-Stress). Each entered the meta-learner as a separate base learner with its own DML residualization step, identical otherwise to the monolithic-MH primary specification. Sub-modality contributions were read both from per-sub-modality standalone ROC-AUC (mean across the 10 outer cross-validation folds) and from permutation importance on the meta-learner inputs. Because the five sub-scales are highly correlated, this decomposition is interpreted at the level of relative ranking and approximate magnitude rather than as identifiable independent effects. The same caution applies a fortiori within the PHQ-9 sub-modality: since the sum score is the exact arithmetic total of the nine items, that block is rank-deficient by construction. Its predictive performance as a sub-modality is unaffected, but no claim is made about which individual item carries the signal, and we do not attempt a somatic versus affective item decomposition, for which competing published item assignments exist.

### 3.4 Transportability validation (within-study)

As a within-study transfer check across the MRI participation gradient, we applied a fitted non-imaging classifier without refitting to the non-MRI NAKO sample (41,402 infected clean-controls participants; PCC-positive prevalence 28.4%). Because the non-MRI arm by construction lacks the six brain-atlas modalities, the transportability assessment used what we term a *Lean Universal Classifier* restricted to four modalities that are available in both arms: baseline mental health, demographics, socioeconomic status, and medical history. The Lean stack was specified before the transfer analysis. It comprises the modalities that need no laboratory, physiological, imaging or cognitive-test measurement and are also recorded in the cohort planned for external replication, with baseline mental health included by design. The rule also required at least 0.02 PR-AUC over the demographics baseline; on the values reported here (Supplementary Table S8) every modality meets that threshold, so the selection rests on availability. The modality set was not adjusted to the results it is tested against. Within its four modalities the Lean stack uses the same variables as the full model, except the item-level PHQ-9 responses and the tobacco variables, which became available only after its feature set had been fixed. The transportability argument therefore does not come to rest on variables whose availability in an external cohort is unknown. The Lean stack was trained under the identical 10 × 5 nested-CV protocol on the MRI cohort and persisted with its per-modality pipelines, DML residualizers, meta-learner and Platt calibrator intact. It was then applied end-to-end to the non-MRI cohort, with each per-modality pipeline receiving the non-MRI confounder frame for orthogonalization but no refit of any component.

The *a priori* success criterion is a conjunctive two one-sided tests (TOST)-style equivalence test on three literature-standard transportability metrics for clinical prediction models (Van Calster et al., 2016): (i) discrimination, target-cohort 95% CI of the ROC-AUC entirely within 0.03 of the source-cohort point estimate; (ii) calibration slope, target CI entirely within [0.85, 1.15]; (iii) calibration-in-the-large, target CI entirely within [−0.05, 0.05]. All three must pass for an overall transportability verdict of Pass, a deliberately conservative conjunction. Discrimination CIs used the same stratified bootstrap protocol as the primary evaluation (1,000 iterations). Calibration CIs were produced by a secondary stratified bootstrap that refits the per-bootstrap calibration regression on the fixed transfer predictions. This conditions on the source-trained pipeline and therefore captures target-cohort resampling variability but not the additional uncertainty that would arise from refitting the source pipeline, consistent with standard transportability practice (Van Calster et al., 2016). As a descriptive transportability profile we additionally report per-modality SHAP-share 95% percentile intervals from 1,000 bootstrap resamples of source and target OoF matrices (same fitted meta-learner). These are reported as transportability characterization only, not as pass/fail, because at the available *N* their intervals become so narrow that any systematic drift is statistically resolvable regardless of practical relevance.

### 3.5 Reporting and transparency

We report this study in accordance with the Transparent Reporting of a multivariable prediction model for Individual Prognosis Or Diagnosis (TRIPOD)+Artificial Intelligence (AI) statement for prediction models developed with regression or machine-learning methods (Collins et al., 2024). Several of its items are answered here in the negative, which we state rather than omit. The study size was not arrived at by a power calculation: it is fixed by data availability, comprising every NAKO neuroimaging participant with a reported SARS-CoV-2 infection and an observable Corona-2 symptom outcome, which yields 2,302 events against ten candidate modalities. Neither the outcome nor the predictors were assessed with blinding to the other, because both were collected as part of the NAKO examination program years apart and independently of this analysis, so the assessment of one could not have been informed by the other. No formal algorithmic-fairness assessment was performed; we report the sex-stratified association analyses of Section 3.3 but no subgroup-specific discrimination or calibration. The model was neither updated nor recalibrated after development; the transportability analysis applied the fitted model to a second cohort without refitting, and the cohort-specific refits reported alongside it are separate models. There is no separate registered protocol for this secondary analysis beyond the approved data application (No. NAKO-882) and the registration of the parent cohort. No patients or members of the public were involved in the design, conduct, or reporting of this study.

### 3.6 Implementation

All analyses were conducted in Python 3.14 using scikit-learn 1.9 (Pedregosa et al., 2011), XGBoost 3.4 (Chen & Guestrin, 2016), pandas 3.0, and NumPy 2.5. The weighted PCS (see Section 2.2) was computed directly from the Corona-2 symptom items without external dependencies. Random seeds were fixed at 42 for all stochastic operations. The analysis code is publicly available at https://github.com/cl445/pcc-prediction-nako.

## 4 Results

### 4.1 Sample characteristics

Table 2 presents participant characteristics within the clean-controls analytic sample, stratified by PCC status. Age was balanced between groups, whereas the PCC group was markedly more female and showed elevated depression and anxiety symptoms on the Corona-2 PHQ-9 and GAD-7. The Corona-2 psychometric scores are reported for descriptive purposes only and are not used as predictors, since temporal co-occurrence with the outcome precludes causal interpretation. Baseline PHQ-9 and GAD-7 (assessed 2014–2019) were also elevated in the PCC group, consistent with a pre-existing mental-health signal rather than concurrent distress alone, and these baseline scores entered the mental-health modality as predictors. Body Mass Index (BMI) and educational attainment differed modestly between groups.

**Table 2:** Baseline characteristics. Participant characteristics by PCC status within the clean-controls analytic sample (*N* = 8,464: 6,160 PCC-negative and 2,304 PCC-positive; infected MRI participants with observable symptom outcome, restricted to PCS-positive cases and declared-symptom-free controls). Cross-validation runs on *N* = 8,461 (Figure 1). Values are mean ± SD for continuous variables and *n* (%) for categorical variables. *p*-values were calculated using Welch’s *t*-test for continuous variables and *χ*^2^ test for categorical variables. ^†^Assessed during Corona-2 follow-up (2022–2023), reported for descriptive purposes only (not used as predictors); all other variables assessed at NAKO baseline (2014–2019). PCC is defined as weighted PCS > 10.75 (Bahmer et al., 2022).

| Variable | PCC– ($n = 6,160$) | PCC+ ($n = 2,304$) | $p$ |
| --- | --- | --- | --- |
| Age, years | 45.3 ± 11.9 | 45.0 ± 11.3 | 0.33 |
| Female sex | 2,442 (39.6%) | 1,258 (54.6%) | <0.001 |
| BMI, kg/m<sup>2</sup> | 25.9 ± 4.3 | 26.3 ± 4.8 | <0.001 |
| BMI category |  |  | <0.001 |
| Underweight (<18.5) | 49 (0.8%) | 27 (1.2%) |  |
| Normal (18.5–24.9) | 2,872 (46.7%) | 1,028 (44.6%) |  |
| Overweight (25–29.9) | 2,325 (37.8%) | 829 (36.0%) |  |
| Obese ($\geq 30$) | 910 (14.8%) | 419 (18.2%) | |
| Education (ISCED) | 4.5 ± 0.9 | 4.4 ± 1.0 | <0.001 |
| Currently employed | 5,517 (89.9%) | 2,078 (90.4%) | 0.10 |
| Hypertension | 1,083 (17.6%) | 454 (19.7%) | 0.03 |
| Cancer history | 247 (4.0%) | 114 (4.9%) | 0.07 |
| PHQ-9 score (0–27)<sup>†</sup> | 3.1 ± 3.4 | 6.9 ± 4.6 | <0.001 |
| GAD-7 score (0–21)<sup>†</sup> | 2.6 ± 3.0 | 5.3 ± 4.1 | <0.001 |
| PHQ-9 baseline (0–27) | 3.1 ± 2.9 | 4.7 ± 3.8 | <0.001 |
| GAD-7 baseline (0–21) | 2.6 ± 2.6 | 3.8 ± 3.2 | <0.001 |

PCC = Post-COVID Condition; BMI = Body Mass Index; ISCED = International Standard Classification of Education; PHQ-9 = Patient Health Questionnaire-9; GAD-7 = Generalized Anxiety Disorder-7.

### 4.2 Overall model performance

The multi-modal meta-learner achieved a ROC-AUC of 0.665 (95% CI, 0.653–0.677) and a PR-AUC of 0.413 (95% CI, 0.392–0.434) for predicting binary PCC status (Table 3), significantly exceeding chance performance (*p* < 0.001, 1,000-iteration meta-permutation test; null-distribution mean PR-AUC = 0.273). Performance was stable across the 10 outer cross-validation folds (SD ≤ 0.032 for both metrics; Supplementary Table S4). The model was well calibrated, with low expected calibration error (ECE = 0.013; Figure 4C) and a calibration slope indistinguishable from 1 (1.030).

**Figure 4:**
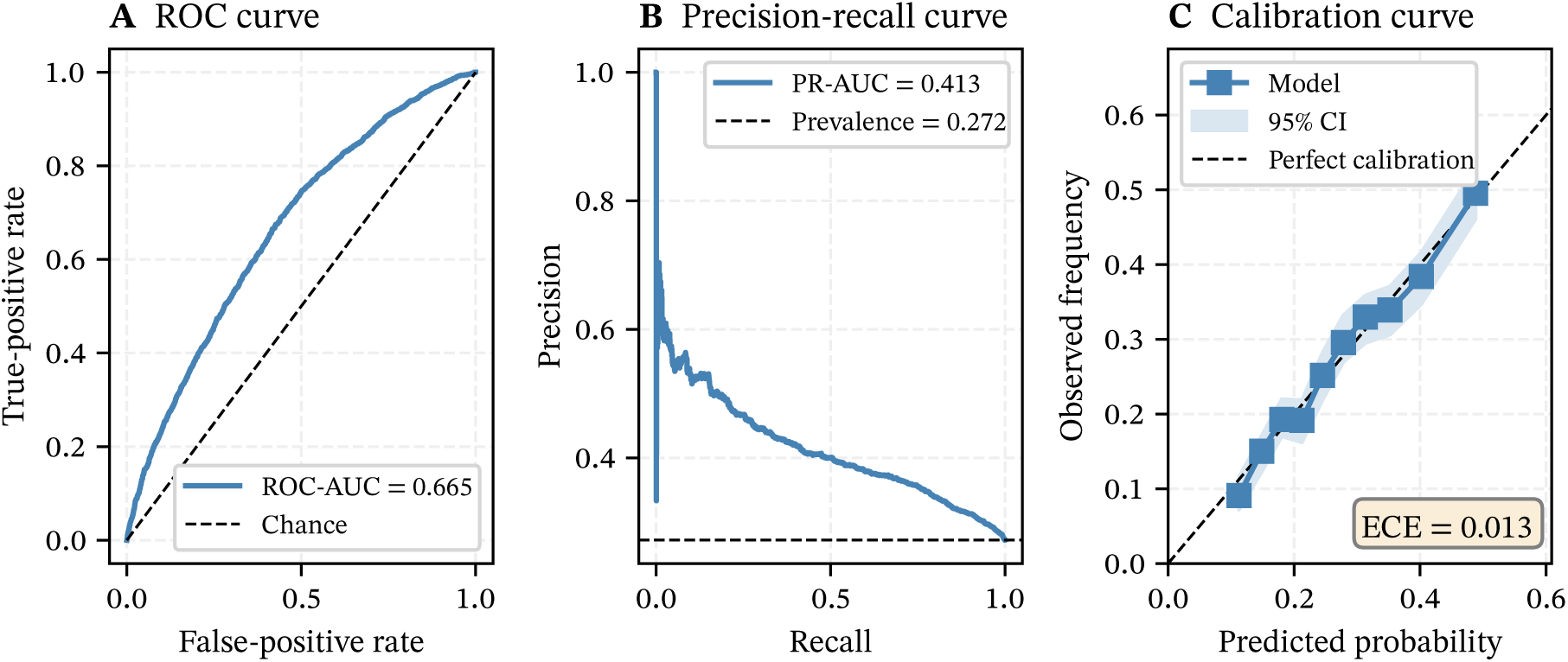
Model evaluation. (A) ROC curve showing sensitivity versus 1-specificity; the dashed diagonal represents chance performance (AUC = 0.5). (B) PR curve showing precision versus recall; the dashed horizontal line represents baseline precision equal to PCC prevalence (27.2%). (C) Calibration curve comparing predicted probabilities to observed frequencies across 10 equal-count (quantile) probability bins, so that each point rests on the same number of participants; the dashed diagonal represents perfect calibration (ECE = 0.013, computed over those same bins on the pooled out-of-fold predictions).

**Table 3:** Model performance metrics. Primary and secondary evaluation metrics for the multi-modal meta-learner predicting binary PCC status (weighted PCS > 10.75) on the clean-controls cross-validation sample (*N* = 8,461; Figure 1). CIs were computed via bootstrap (1,000 iterations), resampled stratified by outcome for ROC-AUC and unstratified for PR-AUC (Section 3.3).

| Metric | Value | 95% CI |
| --- | --- | --- |
| *Discrimination* |  |  |
| ROC-AUC | 0.665 | [0.653, 0.677] |
| PR-AUC<sup>a</sup> | 0.413 | [0.392, 0.434] |
| *Calibration* |  |  |
| Brier Score | 0.185 | – |
| Calibration Slope | 1.030 | – |
| Calibration Intercept | 0.021 | – |
| ECE | 0.013 | – |
| *Statistical Significance* |  |  |
| Permutation $p$-value<sup>b</sup> | <0.001 | – |

<sup>a</sup> Primary evaluation metric, accounting for class imbalance (27.2% PCC prevalence in the clean-controls analytic sample).
<sup>b</sup> Based on 1,000 permutations of PR-AUC; permutation null distribution mean = 0.273.
ROC = Receiver Operating Characteristic; AUC = Area Under the Curve; PR = Precision-Recall; ECE = Expected Calibration Error; CI = Confidence Interval; PCC = Post-COVID Condition.

### 4.3 Modality contributions

Baseline mental health was the strongest single-modality predictor (mean ROC-AUC = 0.640, standard deviation (SD) = 0.022; Table 4), followed by demographics (0.583, SD = 0.019). All six brain MRI sub-modalities clustered near chance (ROC-AUC = 0.496–0.516), with Desikan-Killiany and cerebellar atlases indistinguishable from chance. This near-chance performance is specific to the orthogonalized features: with demographic orthogonalization disabled, the same atlases reached ROC-AUC 0.559–0.570 (Supplementary Table S9), still at or below demographics alone (0.583). The only PCC-predictive content the volumetric features carry is therefore the age/sex axis already supplied by demographics, and the MRI null holds whether or not orthogonalization is applied (the MRI masking contribution is near zero in both regimes). Medical history (0.555, SD = 0.021), SES (0.553, SD = 0.017), and laboratory (0.538, SD = 0.028) provided modest additional discrimination; the remaining biomedical modalities (cardiovascular, lung function, cognitive) and physical activity were similarly uninformative.

**Table 4:** Individual modality performance. Discriminative performance of each modality when used alone, averaged across 10 outer cross-validation folds on the clean-controls analytic sample (*N* = 8,461). Modalities are ranked by mean ROC-AUC. Holm-adjusted *p*-values from Nadeau–Bengio corrected paired tests (Nadeau & Bengio, 2003) compare each modality’s PR-AUC against the mental-health baseline (the strongest standalone modality).

| Modality | ROC-AUC | PR-AUC | SHAP (%) | $p_{\text{Holm}}^b$ |
| --- | --- | --- | --- | --- |
| *Non-imaging modalities* |  |  |  |  |
| Mental health | $0.640 \pm 0.022$ | $0.401 \pm 0.028$ | 38.0 | – |
| Demographics | $0.583 \pm 0.019$ | $0.333 \pm 0.013$ | 27.3 | 0.002 |
| Medical history | $0.555 \pm 0.021$ | $0.319 \pm 0.017$ | 5.8 | 0.002 |
| SES | $0.553 \pm 0.017$ | $0.316 \pm 0.019$ | 8.1 | 0.001 |
| Laboratory | $0.538 \pm 0.028$ | $0.308 \pm 0.022$ | 3.4 | <0.001 |
| Cardiovascular | $0.533 \pm 0.011$ | $0.296 \pm 0.012$ | 4.3 | <0.001 |
| Lung function | $0.524 \pm 0.024$ | $0.287 \pm 0.017$ | 4.5 | <0.001 |
| Cognitive | $0.514 \pm 0.025$ | $0.283 \pm 0.015$ | 1.1 | <0.001 |
| Physical activity | $0.511 \pm 0.014$ | $0.293 \pm 0.016$ | 1.9 | <0.001 |
| *Brain MRI sub-modalities<sup>a</sup>* |  |  |  |  |
| Yeo networks | $0.516 \pm 0.028$ | $0.287 \pm 0.018$ | 0.3 | <0.001 |
| Destrieux | $0.515 \pm 0.024$ | $0.287 \pm 0.025$ | 0.7 | 0.001 |
| Subcortical | $0.514 \pm 0.025$ | $0.282 \pm 0.014$ | 1.2 | <0.001 |
| Julich | $0.510 \pm 0.023$ | $0.277 \pm 0.013$ | 1.2 | <0.001 |
| Desikan-Killiany | $0.498 \pm 0.029$ | $0.276 \pm 0.019$ | 1.8 | <0.001 |
| Cerebellar | $0.496 \pm 0.009$ | $0.270 \pm 0.008$ | 0.3 | <0.001 |

<sup>a</sup> MRI sub-modalities represent different brain parcellation schemes.
<sup>b</sup> Holm-adjusted $p$ -values from Nadeau–Bengio corrected paired $t$ -tests comparing each modality’s PR-AUC against mental health (reference). Every comparison rejected the null of equal performance at $\alpha = 0.05$ ; the largest adjusted $p$ -value across all fourteen was 0.002.
ROC = Receiver Operating Characteristic; AUC = Area Under the Curve; PR = Precision-Recall; SHAP = SHapley Additive exPlanations; MRI = Magnetic Resonance Imaging; SES = Socioeconomic Status.

SHAP-based feature importance ranked baseline mental health first (38.0% of meta-learner signal), followed by demographics (27.3%) and SES (8.1%), with every remaining modality at 5.8% or less (Figure 5). Mental health led by a wide margin on the SHAP and standalone-ROC criteria; the pairwise Nadeau–Bengio PR-AUC comparisons, which are the conservative test of that lead, separated mental health from every one of the other fourteen modalities (Table 4, note b). Modality masking (ΔPR-AUC when replacing each modality’s predictions with NaN in the fitted model) and incremental performance (ΔPR-AUC when adding each modality to a demographics-only baseline) are reported in Supplementary Tables S6 and S8. Adding baseline mental health to the demographics-only baseline (PR-AUC = 0.371) produced by far the largest incremental contribution among all modalities (ΔPR-AUC = 0.122, against 0.059–0.073 for every other modality). The two analyses answer different questions and, where they appear to diverge for the weak modalities, the masking analysis is the more informative. In the incremental analysis the six MRI atlases each raised PR-AUC over the demographics-only baseline by 0.064–0.073, a band that encloses medical history (0.069) and SES (0.068). This apparent contribution is an artifact of the untuned two-input meta-learner evaluated on out-of-fold *training* predictions, under which almost any second modality adds a comparable increment to a single-modality baseline. In the masking analysis, which masks each modality in the *full* model on held-out folds and is the more decisive test of unique contribution, all six atlases stayed at or below ΔPR-AUC = 0.0047, two of them negative. Removing all six atlases together and retraining the meta-learner without them left held-out discrimination unchanged: the full model minus the model without MRI gave ΔROC-AUC = −0.0017 (95% CI, −0.0038 to 0.0004) and ΔPR-AUC = −0.0008 (−0.0052 to 0.0032), so the model without MRI was, if anything, marginally better. Both intervals lie within the equivalence margin of ±0.03 (verdict Pass). Masking the six without retraining lowered performance (0.0132 and 0.0150; Supplementary Table S7), because a meta-learner fitted with six inputs then receives them all missing at once, a pattern it never saw in training; the retrained comparison is the one that measures what MRI adds. In the per-modality masking analysis, baseline mental health contributed substantially (ΔPR-AUC = 0.0722, against 0.0225 for demographics and ≤ 0.0047 for every other modality), whereas every other modality, MRI included, contributed little and to a broadly similar degree.

**Figure 5:**
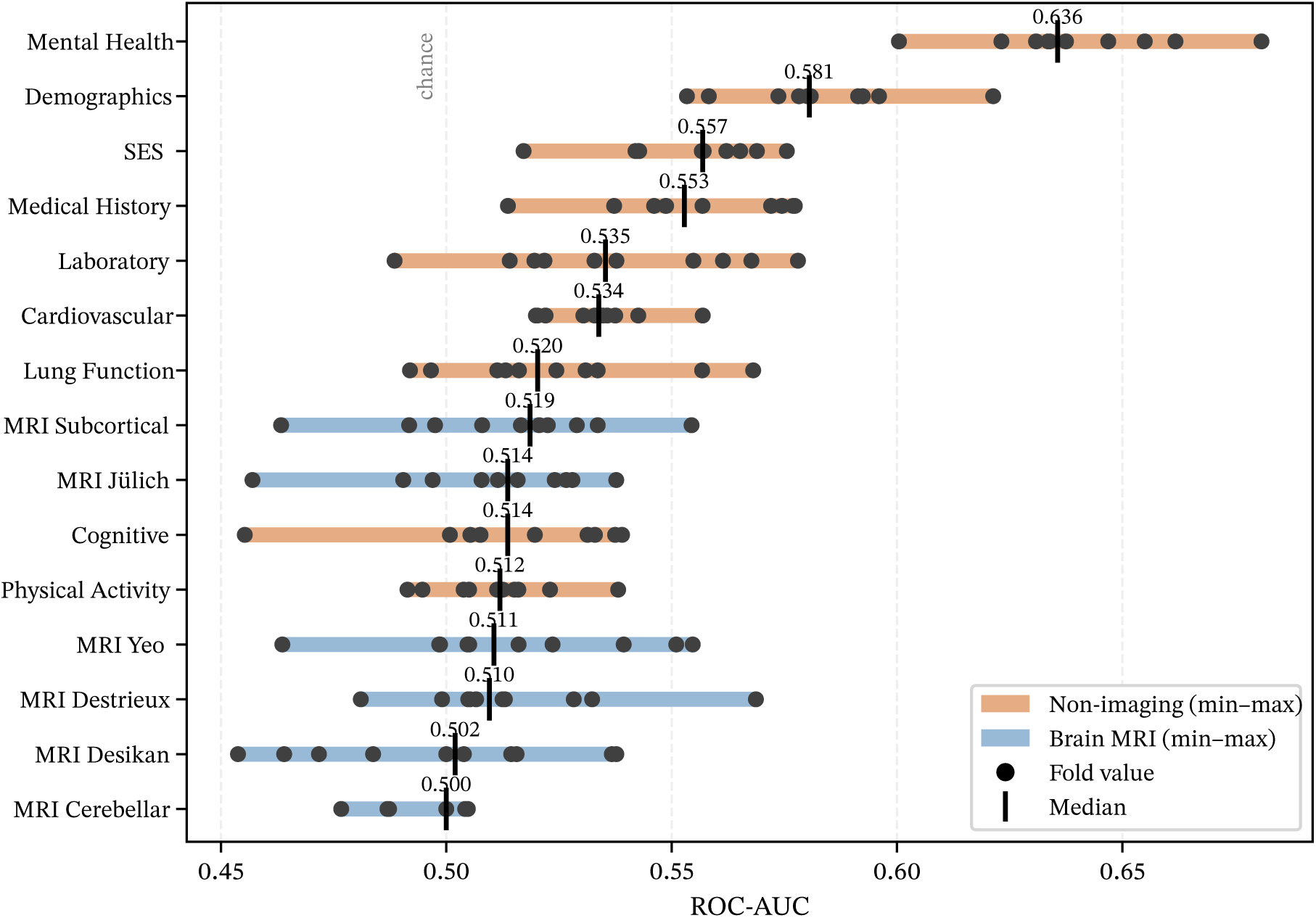
Modality contributions to PCC prediction. Individual modality performance (ROC-AUC) across the 10 outer cross-validation folds. Each modality row shows the per-fold values (black dots), the min–max range (colored bar), and the median (vertical tick). Modalities are colored by type: non-imaging (orange) and brain MRI parcellations (blue). The dashed vertical line indicates chance-level performance (AUC = 0.5). Baseline mental health dominates prediction, followed by demographics, while all MRI sub-modalities and the biomedical modalities (cardiovascular, lung function, cognitive) cluster near chance, with laboratory values slightly above. The corresponding PR-AUC results are shown in Supplementary Figure S1. ROC = Receiver Operating Characteristic; AUC = Area Under the Curve; PR = Precision-Recall; PCC = Post-COVID Condition; MRI = Magnetic Resonance Imaging.

### 4.4 Within-study transportability (non-MRI cohort)

To test whether the predictive structure generalizes beyond the MRI sub-study (which selects younger, healthier, higher-SES participants than the broader NAKO cohort), we applied the fitted Lean Universal Classifier (Section 3.4) to the non-MRI clean-controls cohort (41,402 infected participants; PCC-positive prevalence 28.4%) without any retraining. Source and target cohort baseline characteristics are compared side-by-side in Supplementary Table S11: the non-MRI arm was on average 1.2 years older, had 9.5 percentage points more women, and showed slightly higher cancer history (all standardized mean differences |*d*| < 0.2), consistent with the known MRI participation gradient.

On the MRI cohort, the Lean classifier reached ROC-AUC 0.658 (95% CI, 0.645–0.672), PR-AUC 0.404, calibration slope 1.043, and calibration-in-the-large 0.036. Transferring the same model to the non-MRI cohort yielded ROC-AUC 0.660 (95% CI, 0.654–0.665; Δ = +0.002), calibration slope 1.026 (0.988–1.066), and calibrationin-the-large +0.010 (−0.022 to +0.044). All three pre-specified equivalence sub-tests passed (Supplementary Table S12): the target-cohort 95% CI on ROC-AUC lay within the ±0.03 band, the calibration-slope CI within the [0.85, 1.15] band of Van Calster et al. (2016), and the calibration-intercept CI within [−0.05, 0.05]. The intercept passed with the least room of the three: its upper limit (+0.044) lies close to the band edge, and under a post hoc unstratified bootstrap the pass depends on the random seed, failing for 8 of 20 seeds, and the model-based Wald upper limit (0.051) lies just outside it (Supplementary Table S12 and the text that follows it). The intercept criterion should therefore be read as met narrowly. Expected calibration error (ECE 0.004) and Brier score (0.190 against 0.186 in the source) likewise indicate no material miscalibration on the target. The transfer thus preserves discrimination and calibration with no practically relevant loss: the mental-health-driven predictive signal documented in the MRI sample is not a property of the selected sub-cohort but reproduces in the non-MRI clean-controls sample of the NAKO Corona-2 respondents.

As an additional upper-bound benchmark we refitted the same Lean Universal Classifier directly on the non-MRI cohort under the identical 10 × 5 nested-CV protocol; the cohort-specific refit reached ROC-AUC 0.664 (95% CI, 0.659–0.670), PR-AUC 0.427 (0.418–0.436), calibration slope 1.011, calibration-in-the-large 0.009, and ECE 0.004. The ΔROC-AUC between cohort-specific refit and no-refit transfer is +0.005: the MRI-trained source model extracts nearly all the signal that a non-MRI-trained model extracts from its own data, so refitting on the target cohort gained little.

As a descriptive transportability profile, the SHAP share attributed to baseline mental health remained close to identical between arms: 44.6% in the source (95% CI, 44.3–45.0) versus 44.2% in the target (44.0–44.3). Across all four Lean modalities the modality ranking was preserved, consistent with the performance-equivalence verdict.

As a third NAKO-internal benchmark, refitting the Lean Universal Classifier on the pooled MRI and non-MRI clean-controls union (*N* = 49,865: 8,463 MRI participants, two more than in the primary sample because the Lean stack carries no atlas modality, and 41,402 without MRI) reached ROC-AUC 0.665, indistinguishable from the non-MRI refit (as expected, given that the non-MRI arm dominates the pool at 83% of *N*). Calibration is on par with the other Lean fits (slope 1.009, ECE 0.006), and the modality ranking is consistent with both single-cohort fits.

### 4.5 Hypothesis-aligned neurocognitive subtype

The second-hit hypothesis is framed most directly around neurocognitive sequelae (fatigue, reduced physical capacity, memory, and concentration) as the phenotype most often attributed to central-nervous-system involvement. A pre-specified secondary analysis therefore restricted the PCC outcome to a four-item neurocognitive subtype (endorsement of ≥ 2 of these four items; Section 3.3), yielding an analytic sample of *N* = 7,843 with a prevalence of 21.5%. The clean-controls rule is applied to this outcome in its own right: symptomatic participants with fewer than two of the four items are dropped, while those with two or more count as cases even if their weighted PCS stays at or below 10.75. The sample is therefore not a subset of the primary one, and its case count differs from that in Supplementary Table S5, which applies the subtype definition within the primary sample. This outcome is conceptually narrower than the weighted PCS but more mechanism-aligned: it isolates the exact symptom cluster that a structural cerebral first-hit would be expected to amplify.

Overall discrimination was stronger than for the primary PCC outcome: ROC-AUC = 0.699 (95% CI, 0.685–0.712), PR-AUC = 0.379 (95% CI, 0.358–0.404; permutation *p* < 0.001). PR-AUC was numerically lower than under the primary outcome, reflecting the lower prevalence of the neurocognitive label (21.5% vs. 27.2% in the primary analytic sample) rather than weaker modality separation; discrimination per se, as quantified by ROC-AUC, was higher.

Despite the stronger discrimination, the modality-ranking pattern was preserved (Table 5). Baseline mental health remained the single most informative modality (ROC-AUC = 0.668, SHAP 40.4%), followed by demographics (ROC-AUC = 0.588; SHAP 28.8%). All six brain MRI sub-modalities remained at or near chance (ROC-AUC = 0.498–0.519), with the combined SHAP contribution of all six atlases totaling below 6% of the meta-learner signal. For the outcome most often attributed to central-nervous-system involvement, and therefore the one most likely to be linked to pre-existing volumetric MRI, pre-infection MRI contributes no predictive signal, while pre-infection mental-health phenotype does.

**Table 5:** Individual modality performance: neurocognitive subtype. Discriminative performance of each modality on the neurocognitive subtype outcome (≥ 2 of four items: fatigue, reduced physical capacity, memory, concentration; *N* = 7,843, prevalence 21.5%). Format and conventions as in Table 4. Modalities are ranked by mean ROC-AUC across the 10 outer cross-validation folds.

| Modality | ROC-AUC | PR-AUC | SHAP (%) |
| --- | --- | --- | --- |
| *Non-imaging modalities* |  |  |  |
| Mental Health | $0.668 \pm 0.025$ | $0.366 \pm 0.031$ | 40.4 |
| Demographics | $0.588 \pm 0.034$ | $0.270 \pm 0.024$ | 28.8 |
| Medical History | $0.562 \pm 0.031$ | $0.264 \pm 0.023$ | 6.1 |
| SES | $0.559 \pm 0.029$ | $0.261 \pm 0.021$ | 5.9 |
| Cardiovascular | $0.541 \pm 0.018$ | $0.246 \pm 0.013$ | 5.0 |
| Laboratory | $0.536 \pm 0.025$ | $0.248 \pm 0.019$ | 1.6 |
| Lung Function | $0.527 \pm 0.029$ | $0.231 \pm 0.017$ | 2.5 |
| Cognitive | $0.518 \pm 0.018$ | $0.230 \pm 0.016$ | 1.0 |
| Physical Activity | $0.515 \pm 0.019$ | $0.238 \pm 0.017$ | 3.5 |
| *Brain MRI sub-modalities* |  |  |  |
| Destrieux | $0.519 \pm 0.021$ | $0.237 \pm 0.017$ | 1.4 |
| Desikan-Killiany | $0.510 \pm 0.015$ | $0.227 \pm 0.016$ | 1.0 |
| Yeo Networks | $0.509 \pm 0.030$ | $0.226 \pm 0.022$ | 0.3 |
| Julich | $0.508 \pm 0.022$ | $0.223 \pm 0.018$ | 0.6 |
| Cerebellar | $0.500 \pm 0.014$ | $0.217 \pm 0.009$ | 0.6 |
| Subcortical | $0.498 \pm 0.027$ | $0.224 \pm 0.023$ | 1.4 |

ROC = Receiver Operating Characteristic; AUC = Area Under the Curve; PR = Precision-Recall; SHAP = SHapley Additive exPlanations; MRI = Magnetic Resonance Imaging; SES = Socioeconomic Status.

### 4.6 Mental-health submodality decomposition

To localize which mental-health instruments carry the dominant signal, we refitted the multi-modal pipeline with the monolithic mental-health modality decomposed into five instrument-specific sub-modalities (PHQ-9, GAD-7, MINI major depression, PHQ-Panic, PHQ-Stress) entered as separate base learners (Methods, Section 3.3). Aggregate discrimination was preserved: ROC-AUC = 0.666 (95% CI, 0.653–0.678) versus 0.665 for the monolithic primary, |ΔROC-AUC| < 0.001. Calibration was likewise unaffected (slope 1.03 versus 1.03 in the primary; ECE 0.016 versus 0.013): entering five highly correlated MH base learners into the meta-learner produces no detectable over-confidence.

Per-sub-scale standalone performance and meta-learner permutation importance are reported in Table 6. Three of the five sub-modalities (PHQ-9, PHQ-Stress, and GAD-7) were the top three single-modality predictors over the entire modality field, each above demographics (ROC-AUC = 0.583). PHQ-9 led (mean ROC-AUC 0.632, SD 0.025), followed by PHQ-Stress (0.605, SD 0.027) and GAD-7 (0.602, SD 0.019). MINI-based major-depression diagnosis was weakly above chance (0.530, SD 0.017), and PHQ-Panic carried no signal. Its standalone ROC-AUC was exactly 0.500 with zero across-fold variance (SD 0.000) because its elastic-net base learner shrank all coefficients to zero in every fold, collapsing to a constant predictor rather than discriminating at chance by sampling variability. Meta-learner permutation importance re-ranked these in a similar order: PHQ-9 (0.050), PHQ-Stress (0.009), then MINI (0.004) and GAD-7 (0.004) at the same level, with PHQ-Panic dropping to a negligible contribution. The mental-health signal is thus carried primarily by depressive symptomatology and stress-related complaints, with anxiety as a smaller but separable contributor. Categorical MINI-based major-depression diagnosis adds little beyond the continuous symptom scales, and the PHQ-derived panic items do not contribute predictive signal in this cohort.

**Table 6:**
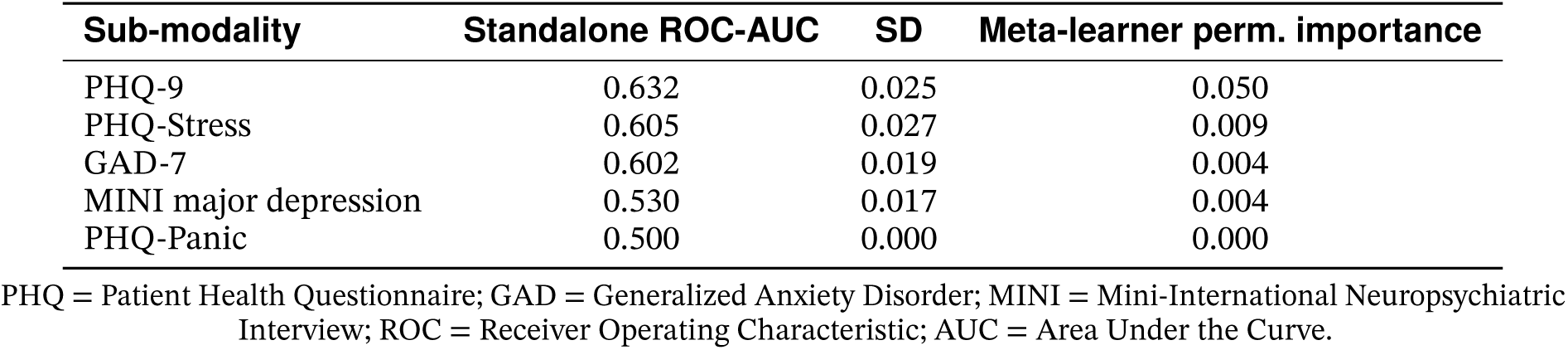
Mental-health sub-scale decomposition. Per-sub-modality standalone discrimination (mean ROC-AUC across 10 outer cross-validation folds; SD across folds) and meta-learner permutation importance, when the monolithic mental-health modality is decomposed into five instrument-specific base learners. Submodalities ranked by standalone ROC-AUC.

| Sub-modality | Standalone ROC-AUC | SD | Meta-learner perm. importance |
| --- | --- | --- | --- |
| PHQ-9 | 0.632 | 0.025 | 0.050 |
| PHQ-Stress | 0.605 | 0.027 | 0.009 |
| GAD-7 | 0.602 | 0.019 | 0.004 |
| MINI major depression | 0.530 | 0.017 | 0.004 |
| PHQ-Panic | 0.500 | 0.000 | 0.000 |

PHQ = Patient Health Questionnaire; GAD = Generalized Anxiety Disorder; MINI = Mini-International Neuropsychiatric Interview; ROC = Receiver Operating Characteristic; AUC = Area Under the Curve.

**Table 7:**
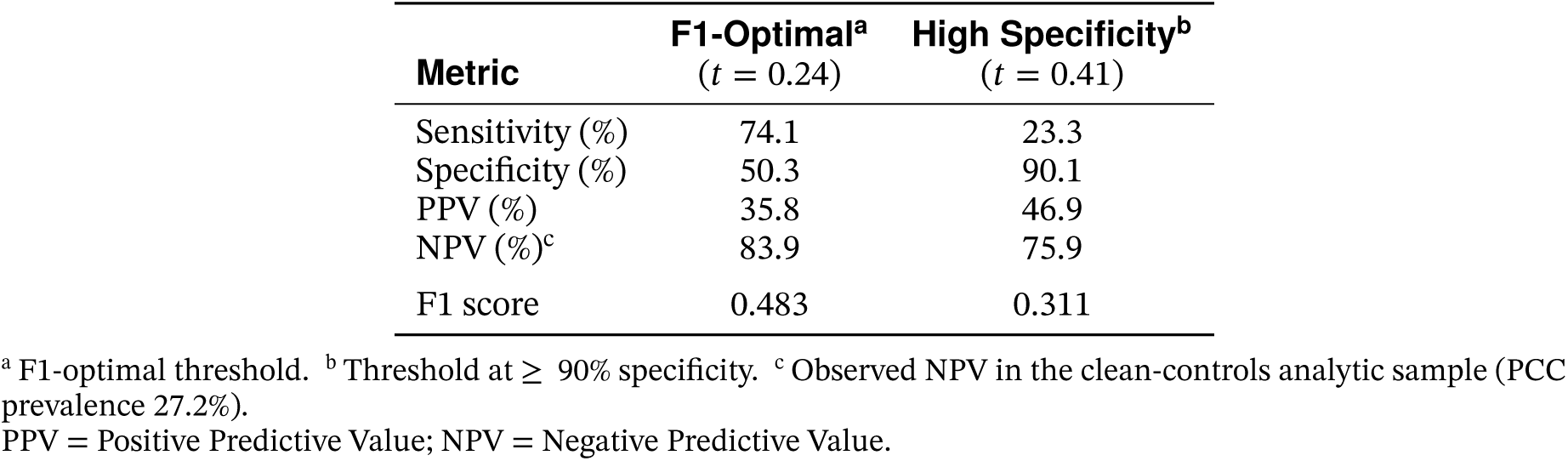
Clinical utility at selected decision thresholds. Performance metrics at the F1-optimal threshold and at a high-specificity threshold suitable for targeted interventions.

### 4.7 Clinical utility

At the optimal F1 threshold of 0.24, the model achieved sensitivity of 74.1%, specificity of 50.3%, and PPV of 35.8% (Table 7). For applications requiring high specificity, a threshold of 0.41 achieved 90.1% specificity with PPV of 46.9%, a 72% relative enrichment over the baseline prevalence of 27.2%. DCA confirmed positive net benefit for threshold probabilities between approximately 8% and 61% (Figure 6).

**Figure 6:**
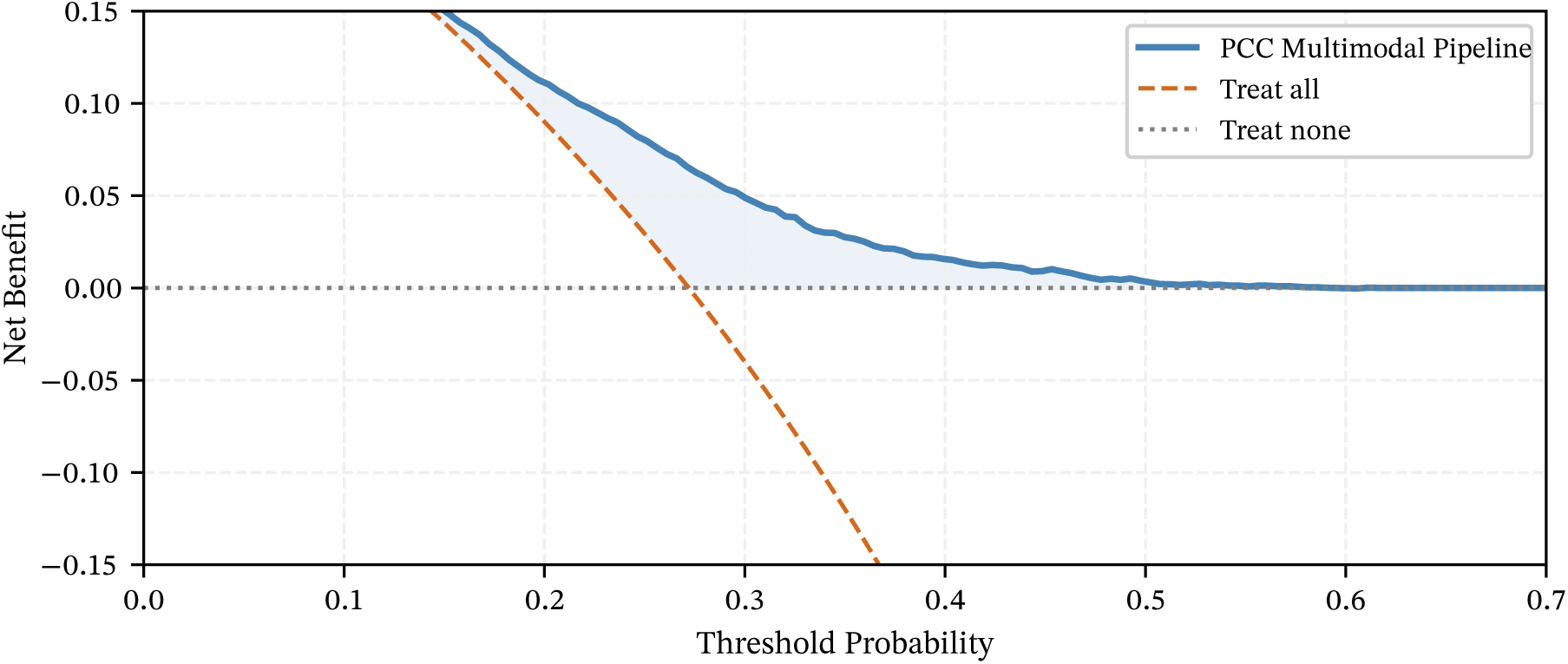
Decision curve analysis. Net benefit of the multi-modal prediction model across threshold probabilities compared to default strategies of treating all patients (orange dashed line) or no patients (dotted line at zero). The model provides a higher net benefit than both default strategies for threshold probabilities between approximately 8% and 61%, indicating clinical utility for risk stratification across a broad operational range.

The Lean Universal Classifier (four non-imaging, non-laboratory modalities; Section 3.4) was evaluated for clinical utility in both NAKO cohorts. Calibration was good in both arms: slope 1.043 (ECE 0.012) on the MRI cohort versus slope 1.011 (ECE 0.004) on the non-MRI cohort, both close to the ideal slope of 1 (Van Calster et al., 2016). At the F1-optimal cut of each arm, sensitivity was 0.795 versus 0.747 and specificity 0.421 versus 0.479; at a high-specificity cut (target ≥ 90% specificity) PPV was 0.465 versus 0.485 and NPV 0.758 versus 0.751. Decision-curve net benefit at the F1-optimal cut was 0.0973 (MRI) versus 0.0941 (non-MRI); at the high-specificity cut 0.0146 versus 0.0164 (Supplementary Figure S2). The non-MRI figures come from a Lean model refitted on that cohort, so they show that the same four modalities support similar clinical utility in both arms; whether the MRI-trained model itself carries over is the question the no-refit transfer in Section 4.4 answers. Both arms are NAKO, so this establishes internal consistency rather than readiness for deployment; the conditions any applied use would have to meet are set out in Section 5.3.

## 5 Discussion

### 5.1 Principal findings

Pre-pandemic baseline mental health, but not structural brain MRI, predicts post-COVID condition years before infection. The multi-modal meta-learner achieves moderate, well-calibrated discrimination (Table 3), and the dual finding (mental health predictive, volumetry not) is preserved under outcome-definition and orthogonalization sensitivity checks, and the mental-health finding also under the transportability check.

#### Baseline mental health is the dominant pre-infection predictor

Baseline mental health is the strongest single-modality predictor. It contributes the largest share of meta-learner feature importance, and it is the modality the full model can least afford to lose: masking it in the fitted model costs 0.072 PR-AUC on held-out folds, about three times the cost of removing demographics and more than ten times that of any remaining modality (Supplementary Table S6). The modality comprises pre-infection scores on the PHQ-9 and the GAD-7, the MINI depression module, and self-reported onset and duration of depressive and anxiety disorders; within it, the PHQ-9 carries most of the signal (Section 4.6). At baseline, PCC-positive participants scored 1.6 points higher on the PHQ-9 and 1.2 points higher on the GAD-7 than PCC-negative participants (*p* < 0.001; Table 2), a difference visible three to eight years before the infection. This pattern replicates prior reports in prospective, online-cohort, and claims-based designs (Bobak et al., 2024; Durstenfeld et al., 2023; Garjani et al., 2022; Greißel et al., 2024; Wang et al., 2022). Whether the association is specific to COVID-19 is taken up in Section 5.2. The three to eight years separating the baseline assessment from infection make reverse causation an implausible account of the association. Pre-infection mental health is therefore a predictor of the outcome rather than an early manifestation of it.

#### Pre-infection structural brain MRI carries no predictive signal

All six brain MRI sub-modalities cluster at or near chance performance (Table 4; ROC-AUC = 0.496–0.516; SHAP ≤ 1.8% each), whether evaluated standalone or within the stacked ensemble, and together the six account for 5.5% of meta-learner feature importance. Removing all six together and retraining the meta-learner without them leaves held-out discrimination unchanged, within the equivalence margin (Supplementary Table S7). If pre-existing volumetric variation constituted a first hit modulating vulnerability to SARS-CoV-2-induced neurological damage, preinfection MRI should carry predictive signal. The present data do not support this operationalization.

This null is a statement about the predictive content of pre-infection volumetry, not about the integrity of the imaging pipeline, including the spatial resolution, image contrast, and sensitivity used in the brain MRI protocol. The same processed atlas features, fed through the same kind of regularized learner *before* orthogonalization, predict participants’ baseline age (combined-atlas *R*^2^ = 0.77, mean absolute error 4.5 years) and sex (ROC-AUC

= 0.984) almost perfectly on the identical analytic sample (Supplementary Table S10). Feature extraction and preprocessing are therefore intact and information-rich, and insensitivity of the MRI pipeline cannot explain the PCC null. The interpretation is instead shaped by how that information is processed for the prediction task. The atlas features are ICV-adjusted and then orthogonalized against age, sex, and recruitment center (Section 3.1). Those are precisely the global-size, age, and sex axes along which a structural “brain reserve” would be expected to vary, so any vulnerability signal aligned with those axes is removed by design. The subsequent PCA retaining 95% of regional-volume variance further summarizes the residual into components in which a weak focal pattern could be diluted. Consistent with this, standalone MRI discrimination rises from chance to ROC-AUC 0.559–0.570 once orthogonalization is disabled (Supplementary Table S9). That is, the only PCC-predictive content the volumetric features carry is the age/sex axis that is already captured by the demographics modality and is not specific to cerebral vulnerability. Any pre-infection volumetric signal is therefore either weak, confined to those non-specific global axes, or carried by dimensions not captured by T1-weighted volumetry. The NAKO T1-weighted volumetric protocol (Bamberg et al., 2015) was designed for whole-body population imaging, not high-resolution neuroanatomy. A signal expressed at finer spatial resolution, in tissue microstructure, or in image contrasts beyond T1-weighted morphometry could thus escape detection here. Higher-resolution or multi-contrast imaging might yet reveal an association between pre-infection brain structure and PCC.

COVID-19 clearly *does* induce structural brain changes: gray-matter thinning and tissue damage after even mild infection (Douaud et al., 2022), oligodendrocyte loss and white-matter myelin dysregulation (Fernández-Castañeda et al., 2022). The absence of pre-infection signal is compatible with these changes arising from the viral and neuroinflammatory insult (Monje & Iwasaki, 2022) rather than reflecting a pre-existing structural vulnerability, although the present data cannot show when they arise. We cannot exclude that vulnerability resides in dimensions not captured by T1-weighted volumetrics (functional connectivity, diffusion-based microstructure, or molecular markers), but pre-infection volumetry, the form in which the structural hypothesis is usually operationalized, carries no predictive signal here.

The two findings constrain each other, and positive controls bracket the MRI null at both relevant levels. The mental-health signal is a positive control at the meta-learner level: the meta-learner picks up a pre-infection signal where one exists. That the MRI null is not a matter of power rests on the joint ablation of the six atlases, whose confidence intervals lie within the equivalence margin (Supplementary Table S7). The near-perfect recovery of age and sex from the same volumetric features is a positive control at the feature-extraction level, ruling out an inert imaging pipeline. Conversely, the MRI null, together with the near-chance performance of the other somatic modalities (see below), argues against the mental-health effect merely reflecting a generic pre-infection baseline-health signal.

#### Demographics and other modalities

Demographics rank second after mental health, but sex carries almost all of this modality’s signal. Univariably, female sex discriminates PCC at ROC-AUC 0.575 (odds ratio (OR) 1.84 [1.67–2.02]), whereas age is at chance (0.491). That chance-level age effect is consistent with the balanced age distribution between groups (Table 2) and with the divergent age effects reported across cohorts (Bai et al., 2022; Subramanian et al., 2022). The female excess risk might involve hormonal, autoimmune, or reporting differences. Baseline mental health itself remained a strong predictor in both sexes when fitted as a sex-stratified composite exposure (adjusted OR 1.89 [1.61–2.21] in women, 2.32 [1.94–2.76] in men), with no significant multiplicative interaction (female-to-male ratio of odds ratios 0.80 [0.63–1.02], *p* = 0.067). The mental-health association is therefore not confined to, or driven by, one sex. SES contributes modestly, while the remaining biomedical modalities (laboratory values, cardiovascular measures, lung function, cognitive function) cluster near chance, providing no evidence that somatic-physiological baseline variation predicts PCC in this cohort.

#### The neurocognitive subtype sharpens both halves of the dual finding

Restricting the outcome to the four-item neurocognitive subtype (fatigue, reduced physical capacity, memory, concentration) yields meaningfully higher discrimination (ROC-AUC = 0.699) than the primary PCC outcome (ROC-AUC = 0.665). This outcome definition is the one most often attributed to central-nervous-system involvement, and therefore the one most likely to be linked to pre-existing cerebral vulnerability. Cognitive fatigue and memory/concentration deficits are the symptom classes for which neuroinflammatory and neuroanatomical mechanisms (Fernández-Castañeda et al., 2022; Monje & Iwasaki, 2022) are most directly implicated.

Two observations follow. First, the improved ROC-AUC shows that baseline phenotyping does carry detectable pre-infection signal for the mechanism-aligned phenotype; pipeline and cohort resolve a stronger signal where the outcome carries one. Second, the modality ranking under the neurocognitive outcome reproduces the ranking under the primary outcome: baseline mental health remains the dominant predictor, while all six MRI sub-modalities remain at or near chance. The neurocognitive outcome is therefore a more mechanism-faithful setting in which to look for a structural signal, and in that setting the null is, if anything, more decisive. For the symptom cluster whose cerebral mediation is most biologically plausible, pre-infection volumetric MRI still adds no signal over and above the psychiatric phenotype.

#### The mental-health signal transports across the MRI participation gradient

MRI participants are younger, healthier, higher-SES and more compliance-affine than the broader Corona-2 cohort (Supplementary Table S3), so any effect estimated within the NAKO neuroimaging sub-study might be confined to this selected arm. We tested this directly with a pre-specified transportability evaluation (Section 3.4). The four-modality Lean Universal Classifier, trained only on the MRI cohort and applied without refit to the non-MRI NAKO cohort, preserved discrimination and calibration. It passed all three pre-specified equivalence bands (ROC-AUC, calibration slope, calibration-in-the-large; Table S12), the last narrowly: under two alternative interval schemes its upper limit touches or crosses the band edge.

The baseline-mental-health SHAP share was likewise near-identical across arms. The mental-health finding therefore holds in the non-MRI clean-controls sample as well and is not a product of the MRI participation gradient.

#### Robustness of the dual finding

Disabling the DML orthogonalization step (Supplementary Table S9) leaves overall discrimination and calibration unchanged (ROC-AUC 0.665 vs. 0.665; calibration slope 1.030 vs. 1.024). The meta-learner SHAP attribution does reallocate, in a way that is itself informative. With orthogonalization off, the predictive credit concentrated in the demographics modality (27.3% → 8.9%) is reabsorbed by the modalities that carry age– and sex-correlated information: baseline mental health (38.0% → 48.7%), medical history, and the MRI atlases (whose summed share rises from 5.5% to roughly 16%). This is what the positive-control finding predicts: the volumetric features encode demographic information that orthogonalization strips out. Baseline mental health remains by far the dominant predictor under both regimes, so the dual finding is preserved.

PHQ-9 and GAD-7 share somatic items with the Corona-2 inventory, so the baseline-MH effect could reflect repeated self-report of the same phenotype. That construct-overlap concern motivated decomposing the 21-item symptom set into five overlap items (fatigue, sleep, concentration, memory, appetite) and 16 mechanism-aligned items. The adjusted baseline-MH odds ratios are indistinguishable between the two outcome families (mechanism-aligned 2.08 vs. overlap 2.26, against 2.06 for the primary PCC outcome), and negative-binomial incidence-rate ratios on symptom counts track this closely. The association is therefore not confined to the outcome items that share content with the depression and anxiety scales. This bounds one form of the concern rather than removing it. A general disposition to report somatic complaints would produce the same flat profile across symptom families, whereas a vulnerability acting through a specific pathophysiology would be expected to produce an uneven one. Flatness alone therefore does not separate the two. Separating them would require decomposing the exposure rather than the outcome, that is, asking which items of the PHQ-9 carry the association and whether the somatic and the cognitive-affective items contribute alike. The present analysis does not do this, and we therefore treat symptom-construct overlap as bounded on the outcome side and open on the exposure side.

Our ROC-AUC of 0.665 sits at the upper end of the 0.60–0.66 range reported for pre-infection PCC prediction models (Doni Jayavelu et al., 2026; Jin et al., 2023; Zang et al., 2024). Post-infection models reach 0.75–0.92 but likely capture acute illness severity rather than pre-existing vulnerability (Butzin-Dozier et al., 2024; Pfaff et al., 2022; Su et al., 2022; Sudre et al., 2021). A Bavarian claims analysis (Greißel et al., 2024) raises an important caveat. Pre-existing psychiatric diagnoses were associated with similar or stronger symptom-persistence risk in non-COVID-19 respiratory-infection controls. The baseline-mental-health effect we observe may therefore index a generalized vulnerability to post-infectious symptom persistence rather than a COVID-19-specific mechanism. Our design cannot adjudicate this distinction because we lack an infection-matched non-COVID-19 control arm.

### 5.2 Strengths and limitations

#### Strengths

Strengths include the large population-based sample, a literature-aligned weighted outcome measure (weighted PCS; Bahmer et al., 2022), nested cross-validation that prevents data leakage, rigorous DML-based confounder control, and calibration and decision-curve evaluation. To our knowledge, no previous study has integrated pre-infection structural brain MRI into a multi-modal PCC prediction framework, or set pre-pandemic neuroimaging against pre-pandemic psychometric assessment as competing candidate predictors in the same participants. The NAKO Level 2 subsample pairs pre-pandemic volumetric MRI with validated psychometric assessment in the same participants, a combination that has been rare. The two candidate operationalizations can therefore be compared jointly instead of in separate cohorts.

#### Outcome circularity

Because our outcome is derived from post-infection symptom endorsement, the symptom items themselves cannot be used as predictor features without circularity. This is a non-trivial restriction, given that post-infection symptom reports typically rank among the strongest predictors in post-hoc PCC models (Pfaff et al., 2022; Sudre et al., 2021). An independent, non-self-report label such as an International Classification of Diseases (ICD)-10 U09.9 diagnosis was in any case unavailable, as NAKO does not link to routine care records. Nor would it be a clean gold standard. U09.9 was introduced only mid-pandemic and is coded sparsely, incompletely, and with long, uneven delay, so an ICD-based outcome would partly track coding behavior and answer a different question. A symptom-based outcome is thus an appropriate primary outcome, though it leaves residual misclassification in both directions and precludes joint modeling of symptom-based features with pre-infection MRI.

#### Design and sample

Three to eight years separated baseline assessment (2014–2019) from infection (2020–2022), possibly too long to capture transient vulnerability states. The neuroimaging subsample (*n* = 19,242, 16.4% of Corona-2 completers) differs demographically from Corona-2 completers without MRI (younger, less often female, and with slightly lower PHQ-9 scores; Supplementary Table S3), although PCC prevalence is indistinguishable between the groups. This limits the generalizability of effect-size estimates to the broader Corona-2 population without compromising internal validity of the hypothesis tests within the neuroimaging subsample. The within-study transfer of the Lean Universal Classifier (Section 3.4) extends the four non-imaging modalities to the non-MRI NAKO arm and therefore corroborates the mental-health finding outside the imaging sub-study. By construction, however, it cannot externally validate the MRI null itself. That null currently rests on the single NAKO Level 2 sample and will require replication in independent pre-pandemic neuroimaging cohorts. PCC heterogeneity across variants, vaccination status, and healthcare settings could additionally obscure domain-specific associations. The outcome also reflects symptom status reported for the 4–12 month post-infection window at the single Corona-2 assessment. It therefore captures whether a participant had developed PCC up to that point rather than whether symptoms persisted afterwards. Cases that resolved or first arose after Corona-2 are not observed.

#### Measurement and statistical resolution

Structural volumetric MRI may miss the relevant vulnerability dimension; the hypothesized first hit could manifest at microstructural (diffusion-derived white-matter integrity), functional (resting-state connectivity), surface-based (cortical thickness or surface area), or molecular levels not assessable from T1-weighted volumetric data (Monje & Iwasaki, 2022). The structural null reported here therefore weighs against the *volumetric* operationalization of pre-infection cerebral vulnerability within one feature-processing pipeline, and only provisionally. The orthogonalization removes the global-size, age, and sex axes on which a structural reserve would vary (above), so a vulnerability aligned with those axes would be undetectable by design. The null likewise does not adjudicate the microstructural, functional, surface-based, or molecular operationalizations, which await dedicated, independent data. We do not stratify the imaging models by acute infection severity, so a structural vulnerability × severity interaction, in which volumetric baseline markers predict PCC only after severe acute insult, would not be detected by the present design. The corresponding interaction for baseline mental health is examined in Supplementary Table S13. The Nadeau–Bengio corrected paired test operates at df = 9 under the 10 × 5 nested cross-validation, which limits the resolution of pairwise modality comparisons when effect sizes are small; the primary ROC and PR discrimination metrics are unaffected. More fundamentally, the modality comparisons and the discrimination and calibration estimates all rest on a single realization of the data-generating process. Nested cross-validation resamples within this one NAKO Level 2 dataset, so it yields estimates that are out-of-sample across folds but still internal to a single cohort draw. The reported cross-fold intervals therefore quantify sampling variability conditional on this cohort rather than the between-dataset variability that only independent replication can establish. The modality ranking, although stable across folds, accordingly awaits confirmation in a separate sample.

#### Causal interpretation

The mental-health signal is temporally pre-infection but correlational: baseline psychiatric phenotype may either reflect a causal vulnerability or index shared upstream factors such as life adversity, chronic inflammation, or reporting style. Interventional evidence, for instance whether pre-infection mental-health care modifies PCC incidence, would be required to anchor this finding causally. We can, however, bound its sensitivity to unmeasured confounding. The age-, sex-, and center-adjusted odds ratio for a composite baseline-MH-positive indicator (PHQ-9 ≥ 10, GAD-7 ≥ 10, or MINI major depression) is 2.06 (1.83–2.32), giving an E-value (VanderWeele & Ding, 2017) of 2.66 (2.42 at the lower CI bound). An unmeasured confounder would thus have to be associated with both baseline mental health and PCC by risk ratios of at least 2.66 each, beyond the measured adjustments, to explain the point estimate away, and of at least 2.42 to move its confidence interval to include the null. This bounds confounded explanations quantitatively without excluding them.

If the association does reflect a biological pathway, candidate mechanisms are available. Depression and anxiety are bidirectionally linked with chronic low-grade systemic inflammation (Beurel et al., 2020), including symptom-specific inflammatory signatures for fatigue-like and neurovegetative features (Milaneschi et al., 2021), and with hypothalamic-pituitary-adrenal axis dysregulation and altered immune reactivity. Any of these could modulate the severity and resolution of post-viral sequelae. The inverse direction of the same axis is consistent with elevated post-COVID-19 rates of mood and anxiety disorders (Taquet et al., 2021) and with the high prevalence of fatigue and cognitive impairment in post-acute sequelae (Ceban et al., 2022). None of this is tested here, and the same literature is equally compatible with both conditions arising from a shared upstream process.

#### Behavioral risk factors

Two behavioral exposures deserve explicit mention, because both are established correlates of depressive symptoms and of general health status and could therefore carry part of the association rather than merely adding to it. Smoking is measured at baseline, enters the multi-modal classifier within the medical-history modality (status, pack-years, duration, lifetime average cigarettes per day), and can also be entered directly into the composite-exposure model. Doing so leaves the association essentially where it was. Adjusting additionally for smoking status and pack-years moves the odds ratio from 2.06 to 2.02 (1.80–2.28), a log-odds shrinkage of 2.2%, and the E-value from 2.66 to 2.62. Tobacco therefore accounts for almost none of the baseline-mental-health effect.

The tobacco terms are themselves informative about why. Neither former (OR 1.00 [0.88–1.14]) nor current smoking (0.96 [0.82–1.12]) predicts PCC once age, sex, and center are accounted for; only cumulative dose does, at 1.010 per pack-year. A weak, dose-dependent predictor that is largely orthogonal to the psychiatric phenotype is not a plausible carrier of an association of this size. The near-absent shrinkage shows that directly.

Alcohol consumption, by contrast, was not available in the approved data set and could be neither modeled nor adjusted for. The Lean Universal Classifier used for the within-study transfer excludes both, since its feature set is fixed in advance across cohorts. Residual confounding through alcohol therefore remains possible, and the E-value above states how strong such a confounder would have to be.

A second concern is that baseline mental health may be a proxy for a persistent psychiatric phenotype that continues into the Corona-2 symptom assessment and inflates self-report rather than indexing a pre-infection vulnerability. A three-wave trajectory adjustment partials out PHQ/GAD-derived mental-health states at Corona-1 (T1, 2020) and Corona-2 (T2, 2022–2023) simultaneously. It shrinks the baseline-T0 odds ratio to 1.38 (1.20–1.58; total log-odds shrinkage 55.4%) but leaves it independently significant alongside a moderate Corona-1 and a dominant Corona-2 effect. A simpler two-wave adjustment for concurrent Corona-2 mental health alone gives the same picture (47.1% shrinkage, direct OR 1.46). Pre-pandemic mental health therefore carries information about post-COVID symptom risk that neither the Corona-1 nor the Corona-2 measurement captures. Short of a randomized intervention, this is as far as the present data can separate a pre-infection-vulnerability interpretation from a persistent-state one. Close to half of the total baseline-MH effect nonetheless remains compatible with a persistent psychiatric state.

This adjustment has two limitations. First, the Corona-2 (T2) mental-health state is measured concurrently with the outcome. If PCC itself provokes depressive and anxiety symptoms, conditioning on T2 therefore partials out a *consequence* of the outcome (a mediator or collider) rather than a confounder. Under that causal structure the trajectory-adjusted OR of 1.38 is a conservative lower bound on the pre-infection effect, not an unbiased estimate of it. Second, and for the same reason, the 55.4% shrinkage cannot be cleanly partitioned into “shared with a persistent trait” versus reverse causation running from PCC to current mood. The supported conclusion is therefore the qualitative one, that a distinct pre-infection (T0) signal survives even this aggressive, partly over-adjusted control, while the magnitude of the surviving effect is bounded below rather than point-identified.

#### Shared self-report method

A third, related concern is that the predictor (baseline PHQ-9/GAD-7) and the outcome (post-infection symptom endorsement) are both self-reported. A stable individual disposition toward symptom over-reporting (negative affectivity or a somatic reporting style) could therefore inflate the association independently of any biological vulnerability. Four observations bear on it, and they do not all point the same way. First, the symptom-content decomposition shows the effect is not confined to the items that share content with the depression and anxiety scales (mechanism-aligned OR 2.08 vs. overlap 2.26). A content-based split, however, addresses construct overlap rather than a *general* reporting style, which by definition acts on all self-report instruments. Second, an objective anchor in the same cohort bounds the concern without settling it. Baseline mental health predicts post-infection loss of smell or taste at essentially the same magnitude as the primary outcome (adjusted OR 2.13 [1.89, 2.40], 1,761 chemosensory cases), and that item is the one PCC symptom NAKO also measures with an instrument. Among participants whose Sniffin’ Sticks screening preceded their reported infection (*n* = 7,227), baseline mental health does not predict measured hyposmia (OR 0.89 [0.66, 1.19]) while predicting self-reported loss of smell in the same participants at 1.92 [1.22, 3.03]. The ratio of the two is 2.12 [1.21, 3.57] on a participant bootstrap, and the same null holds among the never-infected (1.06 [0.95, 1.19]). We therefore read the chemosensory item as a further self-report outcome rather than as evidence against a reporting-style contribution. Two observations point the other way. The measured score predicts the self-report about as strongly as the PHQ-9 does. In the smaller stratum screened after infection, baseline mental health predicts the measured and the self-reported outcome at similar magnitude, which is what convergent olfactory vulnerability would look like (Supplementary Table S15). The present sample cannot separate that reading from a reporting-style one. Third, the baseline-MH odds ratio is unchanged under the alternative mixed-controls outcome that retains sub-threshold symptomatic participants as negatives (OR 2.07 [1.84, 2.33], *N* = 8,960, against 2.05 for the clean-controls outcome fitted on the same participants). The clean-controls contrast, which approaches “endorses any persistent symptom” versus “reports being symptom-free” (Supplementary Table S5), therefore does not itself manufacture the effect. Fourth, self-report was unavoidable for both the exposure and the outcome and will have introduced measurement error in each. To the extent that this error is independent between exposure and outcome, it attenuates the observed association; a shared reporting disposition would not, which is the concern above. A residual general-reporting-style contribution nonetheless cannot be excluded. NAKO’s baseline assessment contains no personality or somatization inventory (e.g. a neuroticism scale or a somatic-symptom screen) independent of the PHQ-9/GAD-7, so we cannot directly adjust for trait negative affectivity. Fully separating a reporting-style artifact from a vulnerability signal would require an outcome ascertained by a different method, such as objective signs or examiner rating, that NAKO does not link. We therefore retain shared-method reporting style as a limitation the present design can bound but not eliminate.

#### What this design cannot identify

Our design establishes that pre-infection mental health is predictive among the infected. That is a necessary condition for the second-hit reading that motivated the comparison: a vulnerability which compounds with infection must at least be predictive in the exposed. It is not the defining interaction, however, and this design cannot identify that interaction. One route to that interaction is closed here: an outcome-identical comparison with non-infected participants is impossible, because the 4–12-month post-COVID symptom items were administered only to the infected. The other, a vulnerability × severity gradient, is estimable, and it points against the dose-response reading rather than for it. On the pooled clean-controls cohort the baseline-mental-health association is undiminished after a mild infection (OR 2.25) and absent among the hospitalized (1.02). That interaction holds on the additive scale as well as the multiplicative one and is therefore not explained by the higher PCC prevalence in that stratum (Supplementary Table S13). Neither repeated infection nor pandemic era shows evidence of modification. We read this as the two routes to PCC being partly distinct: where the acute illness is severe enough to require admission, it supplies the risk on its own. That bounds the present claim to PCC after mild COVID-19 rather than contradicting it. As a coarse surrogate we asked whether baseline mental health predicts *current* (Corona-2) mental-health symptom load more strongly in infected than non-infected participants. It does not (multiplicative interaction OR 0.89 [0.81, 0.97]; within-arm OR 6.32 infected vs. 7.21 non-infected), i.e. the association is, if anything, slightly weaker in the infected, with no sign of infection-driven amplification on this trait-stable outcome. This dovetails with the claims-based observation of Greißel et al. (2024) that pre-existing psychiatric diagnoses also predict symptom persistence after non-COVID-19 respiratory infections. Taken together, the baseline-mental-health signal is better read as indexing a generalized vulnerability to persistent symptom reporting than a COVID-19-specific mechanism. We therefore frame our contribution as the identification and validation of pre-infection predictors of PCC among the infected, and leave the question of what generates the association to designs that can address it.

### 5.3 Clinical implications

The moderate performance precludes individual-level prediction but can support population-level risk stratification through well-calibrated probability estimates. At 90% specificity, the model achieves 46.9% PPV, a 72% relative enrichment over baseline prevalence (27.2%). That is potentially useful for targeting preventive interventions such as structured mental-health follow-up, return-to-activity coaching, or early rehabilitation in primary care. The relative enrichment is substantial, but the absolute gains are small. The decision-curve net benefit at the F1-optimal threshold is about 0.09 (Figure 6), and even at the high-specificity operating point roughly half of flagged individuals remain false positives (PPV 46.9%). The classifier is therefore suited to population-level triage and cohort enrichment (for prevention trials or service planning) rather than to individual diagnosis or standalone clinical decision-making. One further qualification applies specifically to triage. The outcome is defined by self-reported symptoms, and the evidence reviewed in Section 5.2 leaves open that part of the discrimination reflects a disposition to report symptoms rather than the symptom burden itself. To the extent that it does, allocating follow-up on the basis of this model would allocate it by reporting style. That is an argument for validating the model against outcomes ascertained by a different method before any applied use, not against the risk-stratification use case as such. Even this population-level use presupposes access to pre-infection phenotyping that routine care rarely records. The primary contribution is consequently the identification and validation of pre-infection predictors rather than an immediately deployable prediction tool. The transportability evidence reported here is within-study: both arms are NAKO, so it establishes that the signal is not an artifact of the neuroimaging sub-cohort, not that the model travels. External validation in an independent cohort remains a precondition for any applied use.

The null MRI finding does not refute the second-hit hypothesis per se but indicates that if pre-existing cerebral vulnerability contributes to PCC risk, it is not captured by pre-infection structural brain volumes. Functional or diffusion imaging, longitudinal pre-/post-infection designs (Douaud et al., 2022), and neuroinflammatory biomarker integration (Monje & Iwasaki, 2022) may be better suited. The dominance of baseline mental health, conversely, suggests that psychiatric history is a candidate input for post-pandemic risk-stratification tools, to be tested after external validation. It also raises the question of whether PCC prevention and rehabilitation services should triage by baseline psychiatric phenotype rather than by somatic or imaging markers.

### 5.4 Conclusions

Baseline mental health assessed three to eight years before infection is the strongest pre-infection predictor of post-COVID condition among the infected. Whether it acts as a vulnerability that the infection compounds is a separate question, and one this design cannot answer. The defining vulnerability-by-infection interaction is not identifiable here, and what indirect evidence we have points away from infection specificity. In contrast, pre-pandemic structural brain MRI does not improve prediction beyond demographic and psychometric factors, leaving the volumetric operationalization of pre-infection cerebral vulnerability unsupported in this neuroimaging cohort. The remaining biomedical baseline markers add little. These findings redirect future work toward functional and microstructural neuroimaging, toward peri– and post-infection neurobiology, and toward testing, after external validation, whether pre-pandemic psychiatric phenotype improves PCC risk stratification in clinical services. Whether that phenotype is a target or only a marker remains open, and interventional studies are needed to establish whether treating it reduces PCC incidence.

## Data Availability

Data of the German National Cohort (NAKO) are not publicly available due to data protection regulations in accordance with the EU General Data Protection Regulation (GDPR). NAKO data can be obtained via an electronic application portal (www.nako.de/transferhub). The analysis code is publicly available at https://github.com/cl445/pcc-prediction-nako.

## Acknowledgments

We thank all participants of the German National Cohort (NAKO) and the staff of this research initiative. Members and affiliations of the NAKO Investigator Consortium can be accessed via www.nako.de/principal-investigators.

Large language models (e.g., Claude, Anthropic) were used for language editing and drafting support. All content was reviewed and validated by the authors.

## Funding

This project was conducted with data (Application No. NAKO-882) from the German National Cohort (NAKO) (www.nako.de). The NAKO is funded by the Federal Ministry of Research, Technology and Space (BMFTR) [project funding reference numbers: 01ER1301A/B/C, 01ER1511D, 01ER1801A/B/C/D and 01ER2301A/B/C], Federal States of Germany and the Helmholtz Association, the participating universities and the institutes of the Leibniz Association.

This work was supported by the Federal Ministry of Research, Technology and Space (BMFTR) within the RESOLVE-PCC consortium [grant numbers 01EQ2409F and 01EQ2409G].

## Ethics approval and consent

The NAKO is performed with the approval of the relevant local ethics committees and in accordance with national law and with the Declaration of Helsinki of 1975 in its current revised version. Written informed consent was obtained from all participants prior to enrollment, covering the baseline examination, the followup questionnaires and the scientific use of the pseudonymized data. The present secondary analysis was conducted under approved data application NAKO-882 and required no additional participant contact. Details are given in Section 2.1.

## Conflicts of interest

C.L.S. reports speaker fees from Siemens Healthineers, Bayer Healthcare and MSD, and a research grant from Siemens Healthineers. F.B. reports consultancy and speaker fees and an unrestricted research grant from Bayer Healthcare, and an unrestricted research grant from Siemens Healthineers. Neither relationship is connected to the present analysis, which uses previously acquired NAKO imaging data and involves no industry funding, input or review. All other authors declare no competing interests.

## Data and code availability

Data of the NAKO are not publicly available due to data protection regulations in accordance with the EU GDPR. NAKO data can be obtained via an electronic application portal www.nako.de/transferhub. The analysis code is publicly available at https://github.com/cl445/pcc-prediction-nako.

*Trial registration* German National Cohort (NAKO): DRKS00037328

## Supplementary Materials

### Supplementary methods

#### S1. Weighted Post-COVID Syndrome (PCS) Score

The primary PCC outcome is the weighted post-COVID syndrome (PCS) score proposed by Bahmer et al. (2022). The score aggregates 12 symptom complexes, each contributing its weight if any constituent symptom was endorsed during the 4–12 month post-infection window:

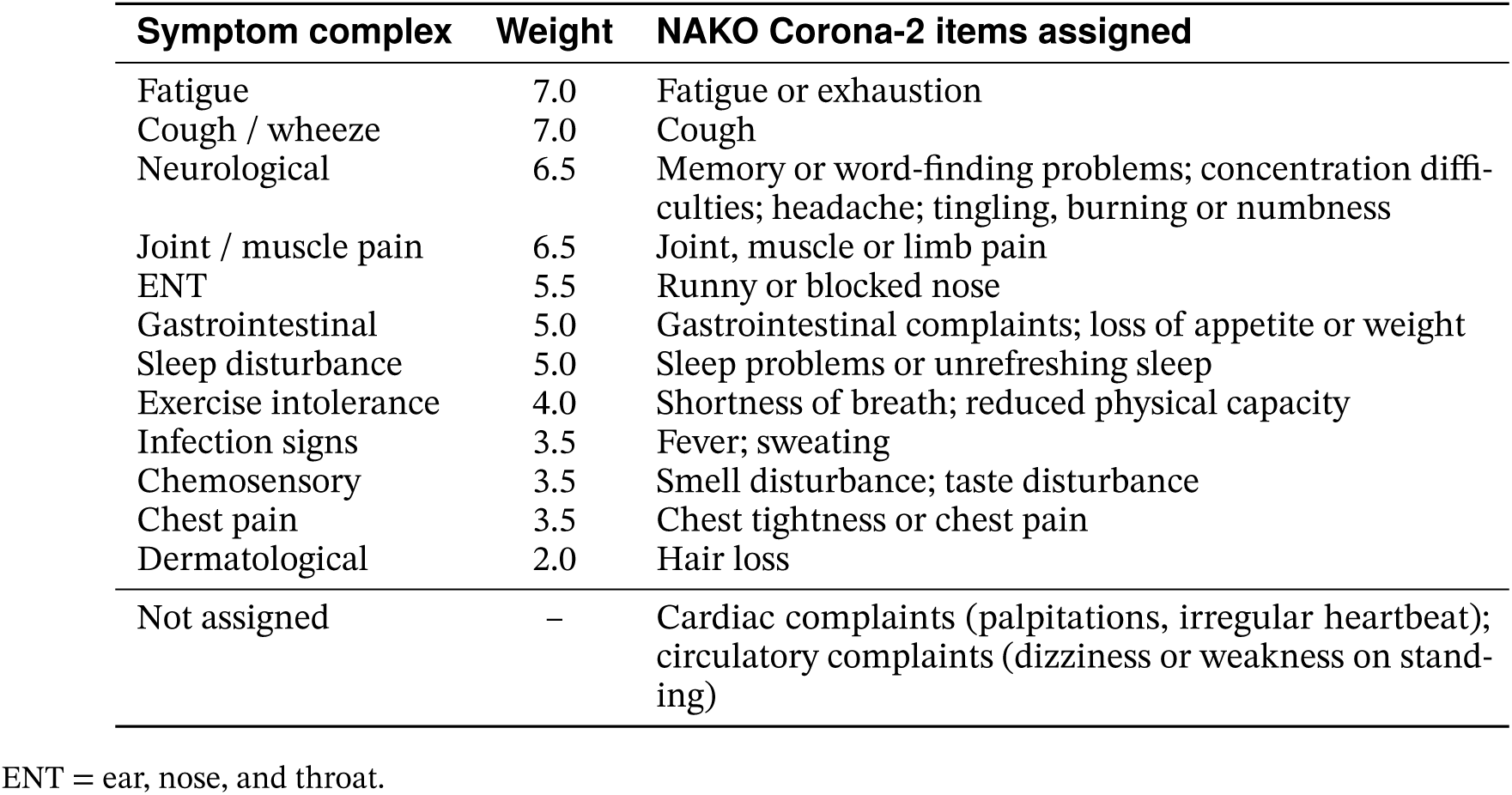

| Symptom complex | Weight | NAKO Corona-2 items assigned |
| --- | --- | --- |
| Fatigue | 7.0 | Fatigue or exhaustion |
| Cough / wheeze | 7.0 | Cough |
| Neurological | 6.5 | Memory or word-finding problems; concentration difficulties; headache; tingling, burning or numbness |
| Joint / muscle pain | 6.5 | Joint, muscle or limb pain |
| ENT | 5.5 | Runny or blocked nose |
| Gastrointestinal | 5.0 | Gastrointestinal complaints; loss of appetite or weight |
| Sleep disturbance | 5.0 | Sleep problems or unrefreshing sleep |
| Exercise intolerance | 4.0 | Shortness of breath; reduced physical capacity |
| Infection signs | 3.5 | Fever; sweating |
| Chemosensory | 3.5 | Smell disturbance; taste disturbance |
| Chest pain | 3.5 | Chest tightness or chest pain |
| Dermatological | 2.0 | Hair loss |
| Not assigned | – | Cardiac complaints (palpitations, irregular heartbeat); circulatory complaints (dizziness or weakness on standing) |

ENT = ear, nose, and throat.

Bahmer et al. (2022) define the complexes by the sub-symptoms of their own interview; the NAKO inventory words its 21 items differently, and we assigned each item to the complex whose sub-symptoms it matches. Sweating and loss of appetite have no exact counterpart and were assigned to the nearest complex. Cardiac and circulatory complaints match none of the twelve and do not enter the score. The total ranges from 0 to 59. Binary PCC status follows the “moderate-or-worse” threshold of > 10.75 reported by Bahmer et al. (2022). The weighted score is computed deterministically from the Corona-2 symptom inventory and requires no external dependencies.

#### S2. Hyperparameter Search Spaces

Meta-learner hyperparameters were tuned via randomized search (50 iterations) with 5-fold cross-validation. Table S1 summarizes the search spaces for each meta-learner variant.

#### S3. Permutation test for statistical significance

Significance of the primary discrimination metric is assessed with a label-permutation test on the fixed out-of-fold predictions. Across 1,000 permutations the class labels are randomly shuffled and PR-AUC is recomputed against the unchanged predictions, yielding a null distribution; the *p*-value is (*r* + 1)/(*B* + 1), where *r* is the number of permuted scores ≥ the observed score and *B* = 1,000. This scheme conditions on the fixed out-of-fold predictions and therefore tests the null that these predictions are label-independent, rather than the broader null of no signal under refit; refit permutation is intractable at the available sample size, but the conditioning is empirically negligible at *N* > 8,000.

#### S4. E-value for unmeasured confounding

To bound the baseline-mental-health association against unmeasured confounding, we fit a logistic regression of the PCC outcome on a dichotomous MH-positive indicator (PHQ-9 sum ≥ 10, GAD-7 sum ≥ 10, or MINI-based major depression) adjusted for age, sex, and study center. The adjusted odds ratio is converted to an approximate risk ratio via the common-outcome correction RR = OR/(1 − *p*_0_ + *p*_0_ ⋅ OR), with *p*_0_ the outcome probability in the unexposed stratum (Zhang & Yu, 1998); this formulation is preferred over the minimax square-<u>root approx</u>imation (VanderWeele, 2020) because *p*_0_ is observed in our sample. The E-value is then RR + √RR(RR − 1) (VanderWeele & Ding, 2017), computed at both the point estimate and the lower 95% CI bound.

**Table S1:** Hyperparameter search spaces. XGBoost meta-learner parameters were sampled via randomized search; base learner parameters were evaluated via grid search using LogisticRegressionCV.

| Component | Parameter | Search Space |
| --- | --- | --- |
| XGBoost meta-learner | Learning rate | {0.01, 0.03, 0.1, 0.2} |
|  | Max depth | {2, 3, 4, 5} |
|  | Subsample ratio | {0.6, 0.8, 1.0} |
|  | Column sample per tree | {0.6, 0.8, 1.0} |
|  | Min child weight | {1, 3, 5} |
|  | Gamma | {0, 0.05, 0.1, 0.2} |
|  | Number of estimators | {100, 200, 400} |
| Base learners<sup>a</sup> | $C$ (inverse regularization) | 4–5 values log-spaced<sup>b</sup> |
|  | L1 ratio | Modality-specific<sup>c</sup> |
| DML nuisance<sup>d</sup> | Number of estimators | 100 |
|  | Max depth | 3 |
|  | Learning rate | 0.1 |
|  | Tree method | hist |

<sup>a</sup> All base learners use LogisticRegressionCV with `scoring=average_precision`, balanced class weights, `max_iter=10,000` (20,000 for SES and medical history), and 3-fold inner CV. Modalities with L1 or elastic net regularization use `solver=saga`; modalities with L2-only regularization (demographics, cardiovascular, lung function) use `solver=lbfgs`. Average precision scoring is used rather than the default log-loss metric, because log-loss favors extreme regularization at low prevalence ( $\approx 27.2\%$ ) and collapses coefficients to zero.
<sup>b</sup> Range by modality group: $[10^{-2}, 10^2]$ for demographics; $[10^{-1}, 10^2]$ for lung function; $[10^{-1}, 10^3]$ for cardiovascular; $[10^{-3}, 10^2]$ for SES, cognitive, physical activity, medical history, laboratory, and mental health; $[10^{-3}, 10^1]$ for MRI atlases. Ranges were fixed per modality group before the analysis and checked once on the neuroimaging subsample (the 19,240 participants with atlas values; imputation and scaling only) to confirm that no modality collapses to a null model; some modalities select $C$ at a grid boundary. The value of $C$ is selected by inner cross-validation inside each outer training fold.
<sup>c</sup> L2 only ( $\ell_1 = 0$ ) for demographics, cardiovascular, and lung function; L2 + L1 ( $\ell_1 \in \{0, 1\}$ ) for SES; elastic net + L1 ( $\ell_1 \in \{0.5, 1\}$ ) for cognitive, physical activity, medical history, laboratory, and mental health; elastic net + L1 ( $\ell_1 \in \{0.5, 1\}$ ) for MRI atlases.
<sup>d</sup> XGBRegressor for DML orthogonalization (Section 3.1). One model per feature, predicting the feature from confounders (age, sex, center). Out-of-fold residuals computed via 5-fold CV.

## Supplementary tables and figures

**Table S2:**
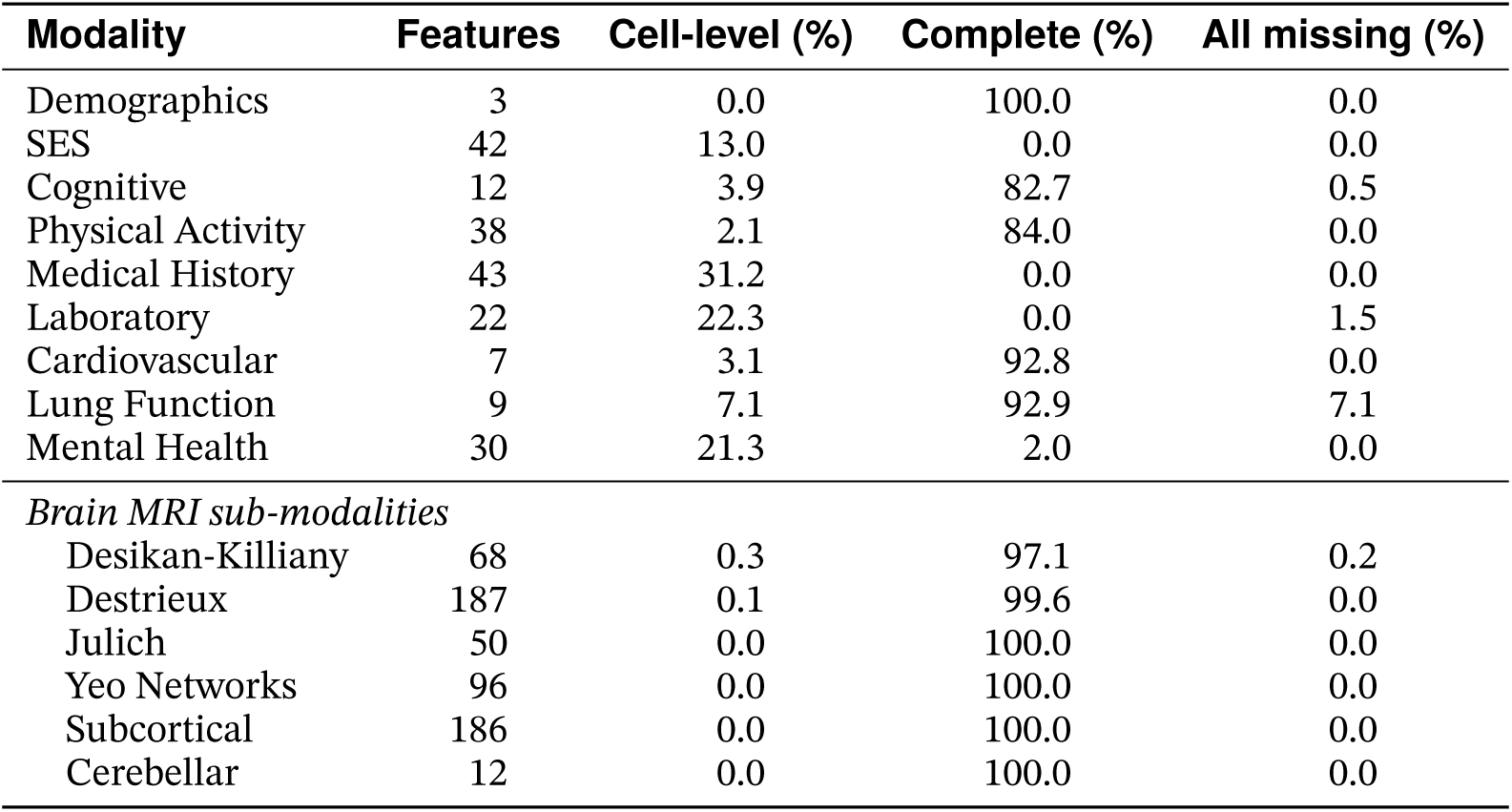
Feature-level missingness by modality. Computed within the clean-controls analytic sample (*N* = 8,464, before the exclusion of three participants missing a whole modality; Figure 1). “Features” denotes the number of raw variables prior to preprocessing (e.g., before one-hot encoding of categorical variables or atlas-specific measure selection); these counts may differ from the analysis-ready feature counts reported in Table 1. “Cell-level” is the percentage of missing values across all feature × participant cells. “Complete” indicates the proportion of participants with no missing values in the modality. “All missing” indicates the proportion of participants missing all features, reflecting systematic non-collection of the entire modality.

**Table S3:**
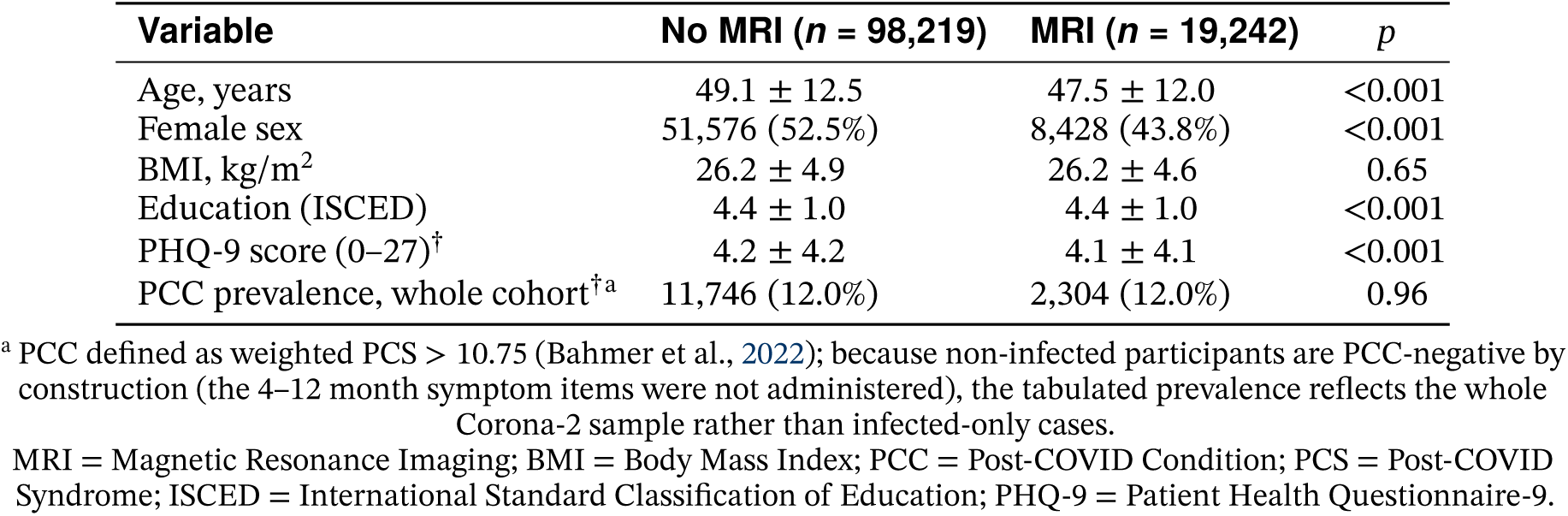
MRI participation gradient. Comparison of *all* Corona-2 participants with and without MRI data (not the infected analytic sample). Values are mean ± SD for continuous variables and *n* (%) for categorical variables. **Note:** the 12.0% PCC prevalence in this table is computed over the whole Corona-2 cohort, in which non-infected participants are PCC-negative by construction; it is therefore *not* comparable to the 27.2% prevalence of the infected clean-controls analytic sample used throughout the rest of the paper (see footnote a). ^†^Assessed during Corona-2 follow-up; all other variables assessed at NAKO baseline.

| Variable | No MRI ($n = 98,219$) | MRI ($n = 19,242$) | $p$ |
| --- | --- | --- | --- |
| Age, years | 49.1 ± 12.5 | 47.5 ± 12.0 | <0.001 |
| Female sex | 51,576 (52.5%) | 8,428 (43.8%) | <0.001 |
| BMI, kg/m<sup>2</sup> | 26.2 ± 4.9 | 26.2 ± 4.6 | 0.65 |
| Education (ISCED) | 4.4 ± 1.0 | 4.4 ± 1.0 | <0.001 |
| PHQ-9 score (0–27)<sup>†</sup> | 4.2 ± 4.2 | 4.1 ± 4.1 | <0.001 |
| PCC prevalence, whole cohort<sup>†a</sup> | 11,746 (12.0%) | 2,304 (12.0%) | 0.96 |

<sup>a</sup> PCC defined as weighted PCS $> 10.75$ (Bahmer et al., 2022); because non-infected participants are PCC-negative by construction (the 4–12 month symptom items were not administered), the tabulated prevalence reflects the whole Corona-2 sample rather than infected-only cases.
MRI = Magnetic Resonance Imaging; BMI = Body Mass Index; PCC = Post-COVID Condition; PCS = Post-COVID Syndrome; ISCED = International Standard Classification of Education; PHQ-9 = Patient Health Questionnaire-9.

**Table S4:** Fold-level performance. Meta-learner prediction performance across 10 outer cross-validation folds. PR-AUC is the primary metric; ROC-AUC is reported for complementary evaluation. Overall calibration metrics (Brier score = 0.185, ECE = 0.013) are computed on the pooled out-of-fold predictions. Low inter-fold variability (across-fold SD 0.032 for PR-AUC and 0.018 for ROC-AUC) indicates stable performance across data splits.

| Fold | PR-AUC | ROC-AUC |
| --- | --- | --- |
| 1 | 0.384 | 0.640 |
| 2 | 0.421 | 0.653 |
| 3 | 0.482 | 0.703 |
| 4 | 0.435 | 0.673 |
| 5 | 0.462 | 0.686 |
| 6 | 0.405 | 0.661 |
| 7 | 0.409 | 0.665 |
| 8 | 0.407 | 0.664 |
| 9 | 0.381 | 0.652 |
| 10 | 0.425 | 0.658 |
| Mean (SD) | 0.421 (0.032) | 0.666 (0.018) |

PR = Precision-Recall; ROC = Receiver Operating Characteristic; AUC = Area Under the Curve; ECE = Expected Calibration Error.

**Table S5:** Outcome-definition agreement. Agreement between PCC outcome definitions within the clean-controls analytic sample (*N* = 8,461). The primary outcome is the weighted PCS (> 10.75; Bahmer et al., 2022). A hypothesis-aligned neurocognitive subtype (endorsement of at least two of four items spanning fatigue, reduced physical capacity, memory problems, and concentration problems) and the PCS severity cut-off (> 26.25) are shown as sensitivity operationalizations. Cohen’s *κ* quantifies agreement beyond chance versus the primary definition. A simple ≥ 1-symptom count (Diexer et al., 2025) is not tabulated because it collapses to the primary definition by construction of the clean-controls arm (controls are explicitly declared symptom-free, so every primary-positive participant is a count-positive and vice versa).

| Definition | PCC+ ($n$) | Prevalence (%) | $\kappa$ vs. primary |
| --- | --- | --- | --- |
| Weighted PCS ($> 10.75$) | 2,302 | 27.2 | – |
| Neurocognitive subtype ($\geq 2$ of 4) | 1,648 | 19.5 | 0.786 |
| PCS severity ($> 26.25$) | 1,219 | 14.4 | 0.621 |

PCC = Post-COVID Condition; PCS = Post-COVID Syndrome.

**Table S6:** Modality masking (held-out). Performance decrease (Δ) when each modality’s base-learner predictions are set to missing and the fold’s own meta-learner re-scores its held-out participants, so that every masked prediction comes from a meta-learner that never saw that participant during training. The full-model reference is therefore the held-out performance reported in the main text (Table 3: ROC-AUC = 0.665, PR-AUC = 0.413). Larger Δ indicates greater contribution to the full model. Because the meta-learner routes missing inputs to a learned default direction rather than to an imputed value, masking measures a modality’s contribution under the model’s learned handling of its absence. It is not a comparison against a model retrained without that modality; Table S7 provides that comparison for the six MRI atlases jointly.

| Modality | ΔROC-AUC | ΔPR-AUC |
| --- | --- | --- |
| Mental Health | 0.0681 | 0.0722 |
| Demographics | 0.0213 | 0.0225 |
| Subcortical | 0.0022 | 0.0047 |
| Physical Activity | 0.0015 | 0.0036 |
| SES | 0.0021 | 0.0030 |
| Cerebellar | 0.0009 | 0.0028 |
| Julich | 0.0017 | 0.0012 |
| Cognitive | 0.0003 | 0.0010 |
| Medical History | 0.0014 | 0.0007 |
| Cardiovascular | 0.0016 | 0.0005 |
| Desikan-Killiany | 0.0003 | 0.0003 |
| Laboratory | 0.0009 | 0.0001 |
| Destrieux | 0.0007 | -0.0002 |
| Lung Function | -0.0002 | -0.0006 |
| Yeo Networks | 0.0013 | -0.0015 |

ROC = Receiver Operating Characteristic; AUC = Area Under the Curve; PR = Precision-Recall; OoF = Out-of-Fold; SES = Socioeconomic Status.

**Table S7:**
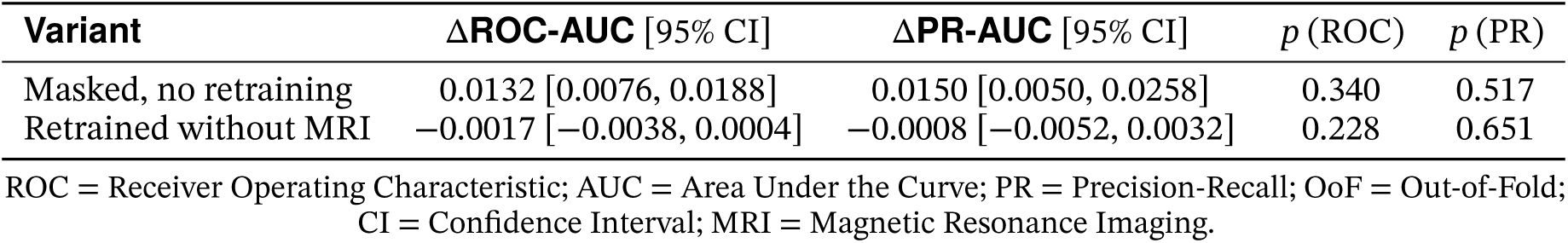
Joint masking and ablation of the six MRI atlases (held-out). Within each outer fold all six atlases were removed together, either by setting their base-learner predictions to missing under the fold’s own meta-learner (as in Table S6) or by retraining the fold’s meta-learner on the training-fold OoF predictions of the nine remaining modalities with the identical hyperparameter search and seed; the held-out fold was scored in both cases and folds were concatenated as for the primary metrics. Δ is the full model minus the model without the atlases, so a positive value is a loss without MRI. 95% CIs from a paired participant bootstrap (1,000 iterations; stratified for ROC-AUC, unstratified for PR-AUC); *p*-values from Nadeau–Bengio corrected paired tests over the 10 per-fold differences. The criterion, fixed before the run but after the per-atlas MRI null was known, is equivalence within ±0.03 on both metrics for the retrained variant (verdict Pass); the masked variant is shown as the contrast between masking and removal.

**Table S8:** Incremental performance. ΔPR-AUC when adding each modality to a demographics-only baseline (demographics PR-AUC = 0.371), computed on OoF training predictions with a two-input meta-learner at default hyperparameters. Under these conditions almost any added modality yields a similar increment, so the increment does not measure a modality’s unique contribution; the held-out masking analysis (Table S6) does.

| Modality | PR-AUC | ΔPR-AUC |
| --- | --- | --- |
| Mental Health | 0.493 | 0.1222 |
| Destrieux | 0.444 | 0.0730 |
| Julich | 0.443 | 0.0723 |
| Cardiovascular | 0.443 | 0.0719 |
| Desikan-Killiany | 0.443 | 0.0716 |
| Yeo Networks | 0.442 | 0.0706 |
| Physical Activity | 0.440 | 0.0691 |
| Medical History | 0.440 | 0.0691 |
| Subcortical | 0.439 | 0.0678 |
| SES | 0.439 | 0.0677 |
| Laboratory | 0.438 | 0.0675 |
| Cerebellar | 0.435 | 0.0644 |
| Cognitive | 0.431 | 0.0603 |
| Lung Function | 0.430 | 0.0593 |

PR = Precision-Recall; AUC = Area Under the Curve; OoF = Out-of-Fold; SES = Socioeconomic Status; MRI = Magnetic Resonance Imaging.

**Figure S1:**
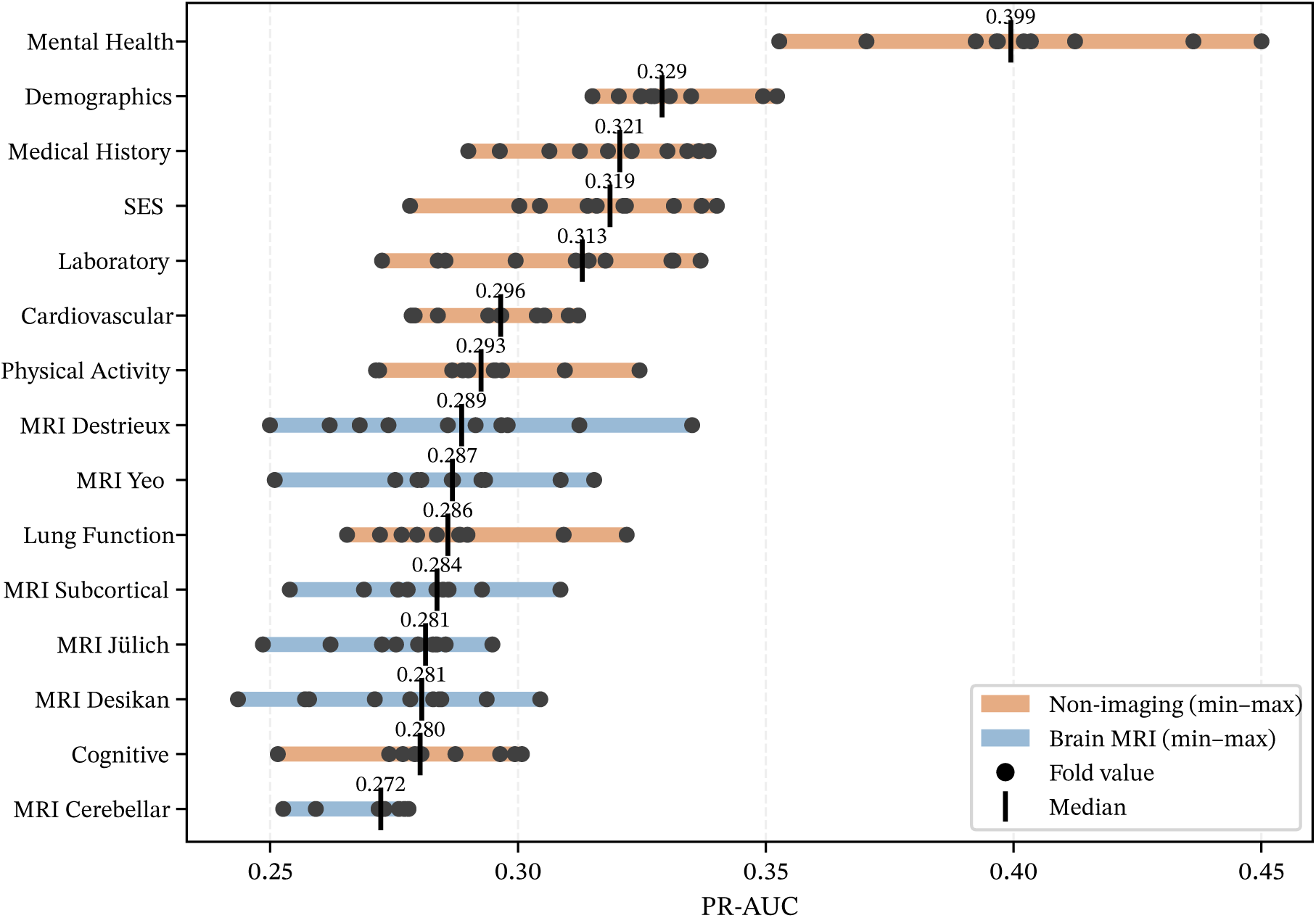
Modality contributions to PCC prediction (PR-AUC). Individual modality performance across 10 outer cross-validation folds. This complements the ROC-AUC results shown in the main text (Figure 5). Modality rankings are consistent across both metrics: baseline mental health leads (median PR-AUC = 0.399), followed by demographics (0.329), while the brain MRI sub-modalities cluster near the baseline prevalence rate (27.2%) and laboratory values sit slightly above them. “Laboratory” is the 19-feature panel of NAKO baseline blood parameters listed in Table 1 (lipids, glucose and HbA1c, CRP, liver, kidney and thyroid function, electrolytes, and blood count). PCC = Post-COVID Condition; PR = Precision-Recall; AUC = Area Under the Curve; ROC = Receiver Operating Characteristic; MRI = Magnetic Resonance Imaging.

**Table S9:** Sensitivity to DML orthogonalization. The primary analysis uses DML-based orthogonalization of each modality’s features against the demographic confounders (age, sex and study center) before the base learner. The sensitivity analysis re-runs the full 10 × 5 nested cross-validation on the clean-controls weighted PCS outcome with orthogonalization disabled. Overall discrimination and calibration are unchanged, and baseline mental health remains the dominant modality in both regimes; the SHAP attribution reallocates in an interpretable way (see note), confirming that the dual finding (baseline mental health dominant, structural MRI near chance) is not an artifact of the orthogonalization step. The bottom block reports each MRI atlas’s standalone ROC-AUC: at chance under the primary DML design (0.496–0.516) but rising to 0.559–0.570 once orthogonalization is disabled, because the un-orthogonalized volumetric features regain the age/sex-correlated signal documented by the positive control (Table S10).

| Outcome / modality | Primary (DML) | Without DML |
| --- | --- | --- |
| *Overall discrimination* |  |  |
| ROC-AUC (95% CI) | 0.665 | 0.665 |
| PR-AUC (95% CI) | 0.413 | 0.417 |
| Calibration slope | 1.030 | 1.024 |
| ECE | 0.013 | 0.013 |
| *Modality SHAP (% of mean \|SHAP\|)* |  |  |
| Mental Health | 38.0 | 48.7 |
| Demographics | 27.3 | 8.9 |
| SES | 8.1 | 6.4 |
| Laboratory | 3.4 | 1.9 |
| Cardiovascular | 4.3 | 5.6 |
| Medical History | 5.8 | 6.7 |
| MRI (Desikan-Killiany) | 1.8 | 6.1 |
| Cognitive | 1.1 | 1.4 |
| MRI (Cerebellar) | 0.3 | 4.5 |
| *Standalone MRI-atlas ROC-AUC (10-fold mean)* |  |  |
| Desikan-Killiany | 0.498 | 0.570 |
| Destrieux | 0.515 | 0.566 |
| Julich | 0.510 | 0.564 |
| Yeo Networks | 0.516 | 0.563 |
| Subcortical | 0.514 | 0.562 |
| Cerebellar | 0.496 | 0.559 |

DML = Double Machine Learning; ROC = Receiver Operating Characteristic; AUC = Area Under the Curve; PR = Precision-Recall; ECE = Expected Calibration Error; SHAP = SHapley Additive exPlanations; CI = Confidence Interval; SES = Socioeconomic Status; MRI = Magnetic Resonance Imaging.

**Table S10:**
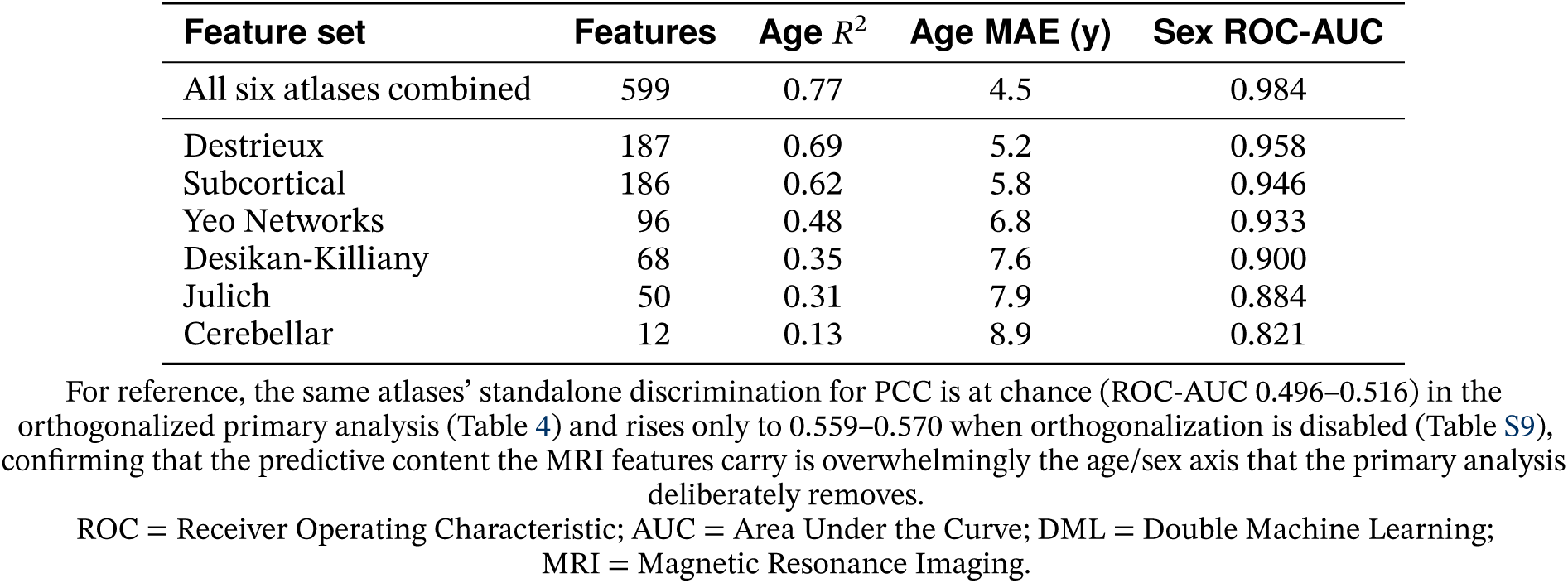
MRI feature-extraction positive control. Out-of-fold (10-fold) prediction of pre-infection baseline *age* (RidgeCV regression; coefficient of determination *R*^2^ and mean absolute error) and *sex* (L2-penalized logistic regression with balanced class weights; ROC-AUC) from the atlas regional-volume features used by the main pipeline, on the identical clean-controls MRI analytic sample (*N* = 8,461), *before* the DML orthogonalization against age, sex, and center. Feature counts are the atlas volumes only and match Table 1; the intracranial-volume covariate used for ICV adjustment is excluded, so the control reflects the regional volumes themselves rather than raw head size. High *R*^2^ and near-perfect sex discrimination establish that the MRI feature extraction and preprocessing deliver intact, information-rich signal, so insensitivity of the imaging pipeline cannot explain the near-chance PCC performance of the same features in the orthogonalized primary analysis. Atlases are ranked by age *R*^2^.

| Feature set | Features | Age $R^2$ | Age MAE (y) | Sex ROC-AUC |
| --- | --- | --- | --- | --- |
| All six atlases combined | 599 | 0.77 | 4.5 | 0.984 |
| Destrieux | 187 | 0.69 | 5.2 | 0.958 |
| Subcortical | 186 | 0.62 | 5.8 | 0.946 |
| Yeo Networks | 96 | 0.48 | 6.8 | 0.933 |
| Desikan-Killiany | 68 | 0.35 | 7.6 | 0.900 |
| Julich | 50 | 0.31 | 7.9 | 0.884 |
| Cerebellar | 12 | 0.13 | 8.9 | 0.821 |

ROC = Receiver Operating Characteristic; AUC = Area Under the Curve; DML = Double Machine Learning; MRI = Magnetic Resonance Imaging.

## Within-study transportability

The side-by-side baseline-characteristics comparison between the MRI and non-MRI clean-controls analytic samples is shown in Table S11. All standardized mean differences are |*d*| < 0.2, so the known MRI participation gradient is small in magnitude despite the very small *p*-values driven by the ∼50,000 combined sample size.

**Table S11:** Baseline characteristics of the MRI and non-MRI clean-controls analytic samples. Counts are the descriptive analytic samples (MRI *n* = 8,464, non-MRI *n* = 41,412), before the drop of participants missing a whole modality (MRI: one mental health, two Destrieux; non-MRI: ten mental health) that yields the cross-validation samples (*N* = 8,461 and 41,402) used for the model performance metrics. Continuous variables reported as mean (SD); binary variables as *n* (%). SMD = standardized mean difference (Cohen’s *d* for continuous, standardized proportion difference for binary). *p*-values from Welch *t*-test (continuous) or *χ*^2^-test (binary / categorical); at the sample sizes shown, SMD is the primary effect-size summary. Hypertension is a physician diagnosis reported at the baseline interview. The MINI was administered to part of the cohort, and the major-depression diagnosis was made only in participants who screened positive on it; its percentages therefore refer to those participants (MRI *n* = 1,698, non-MRI *n* = 1,545), not to the whole sample.

| Variable | MRI ($n = 8,464$) | Non-MRI ($n = 41,412$) | SMD | $p$ |
| --- | --- | --- | --- | --- |
| Age, years | 45.2 (11.7) | 46.4 (12.3) | -0.10 | <0.001 |
| Female sex | 3,700 (43.7%) | 22,023 (53.2%) | -0.19 | <0.001 |
| BMI, kg/m<sup>2</sup> | 26.0 (4.5) | 25.9 (4.7) | 0.02 | 0.044 |
| BMI category |  |  |  | <0.001 |
| Underweight (<18.5) | 76 (0.9%) | 579 (1.4%) |  |  |
| Normal (18.5–24.9) | 3,900 (46.1%) | 19,821 (48.0%) |  |  |
| Overweight (25–29.9) | 3,154 (37.3%) | 14,269 (34.6%) |  |  |
| Obese ($\geq 30$) | 1,329 (15.7%) | 6,601 (16.0%) | | |
| Education (ISCED) | 4.5 (1.0) | 4.4 (1.0) | 0.06 | <0.001 |
| Currently employed | 7,595 (90.1%) | 35,712 (86.8%) | 0.10 | <0.001 |
| Hypertension | 1,537 (18.2%) | 8,367 (20.3%) | -0.05 | <0.001 |
| Cancer history | 361 (4.3%) | 2,405 (5.8%) | -0.07 | <0.001 |
| Number of symptoms<sup>†</sup> | 1.9 (3.6) | 2.0 (3.7) | -0.04 | 0.002 |
| PHQ-9 score (0–27)<sup>†</sup> | 4.1 (4.1) | 4.3 (4.1) | -0.04 | <0.001 |
| GAD-7 score (0–21)<sup>†</sup> | 3.3 (3.6) | 3.5 (3.6) | -0.06 | <0.001 |
| PHQ-9 baseline (0–27) | 3.5 (3.3) | 3.7 (3.4) | -0.05 | <0.001 |
| GAD-7 baseline (0–21) | 2.9 (2.8) | 3.1 (3.0) | -0.05 | <0.001 |
| MINI major depression (baseline) | 1,236 (72.8%) | 1,154 (74.7%) | -0.04 | 0.235 |

<sup>†</sup> Assessed at the Corona-2 follow-up (2022–2023); all other rows at the NAKO baseline (2014–2019). MRI = Magnetic Resonance Imaging; BMI = Body Mass Index; ISCED = International Standard Classification of Education; SMD = standardized mean difference; PHQ-9 = Patient Health Questionnaire-9; GAD-7 = Generalized Anxiety Disorder 7-item scale; MINI = Mini-International Neuropsychiatric Interview.

The Lean Universal Classifier, trained on the MRI cohort and applied without refit to the non-MRI cohort, meets the pre-specified TOST performance-equivalence criterion on all three transportability metrics (ROC-AUC difference, calibration slope and calibration-in-the-large; Table S12 and Section 4.4). Bands follow established clinical-prediction-model transportability thresholds: ±0.03 for ROC-AUC difference (clinical non-inferiority margin), [0.85, 1.15] for calibration slope (acceptable-calibration band per Van Calster et al. 2016), and [−0.05, 0.05] for calibration-in-the-large.

**Table S12:**
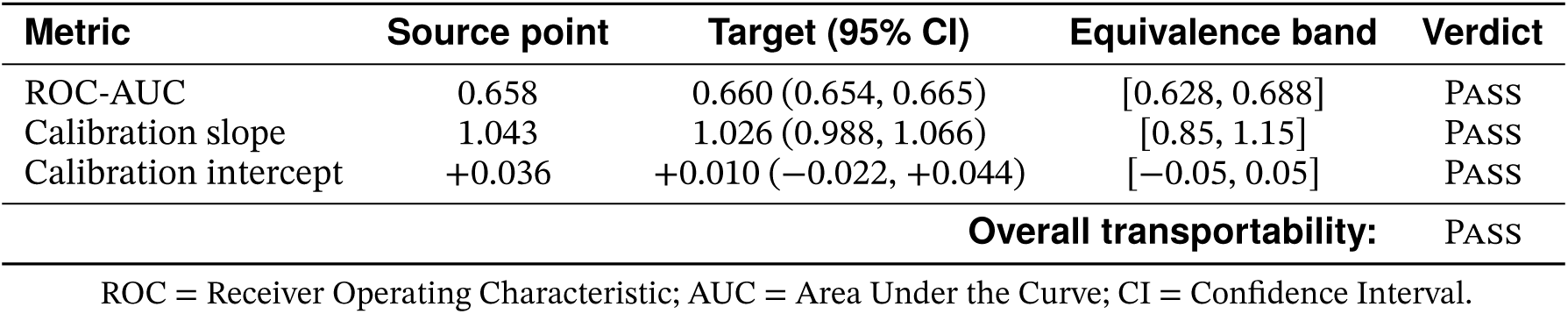
Within-study transportability performance-equivalence panel. Target-cohort 95% CI must lie entirely within the pre-specified equivalence band for PASS. Bands follow literature-standard clinical-prediction-model transportability thresholds (Van Calster et al., 2016).

| Metric | Source point | Target (95% CI) | Equivalence band | Verdict |
| --- | --- | --- | --- | --- |
| ROC-AUC | 0.658 | 0.660 (0.654, 0.665) | [0.628, 0.688] | PASS |
| Calibration slope | 1.043 | 1.026 (0.988, 1.066) | [0.85, 1.15] | PASS |
| Calibration intercept | +0.036 | +0.010 (-0.022, +0.044) | [-0.05, 0.05] | PASS |
| Overall transportability: |  |  |  | PASS |

ROC = Receiver Operating Characteristic; AUC = Area Under the Curve; CI = Confidence Interval.

Of the three criteria, calibration-in-the-large passes with the least room, and it is the one whose pass depends on the interval scheme. The pre-specified calibration interval resamples stratified by outcome, which holds the prevalence fixed, whereas calibration-in-the-large is the quantity most sensitive to prevalence; as a post hoc sensitivity check we therefore also resampled without stratification and computed the model-based Wald interval. The Wald upper limit is 0.051, just outside the band. The unstratified upper limit is 0.049 at the pre-specified bootstrap seed, but it moves with the seed by about as much as the remaining margin: over 20 seeds it averages 0.050 (SD 0.002) and exceeds 0.05 in 8 of them. The intercept criterion is therefore met under the pre-specified scheme but not under the Wald interval and not robustly under the unstratified bootstrap; it is met narrowly.

## Lean Universal Classifier: clinical utility across cohorts

**Figure S2:**
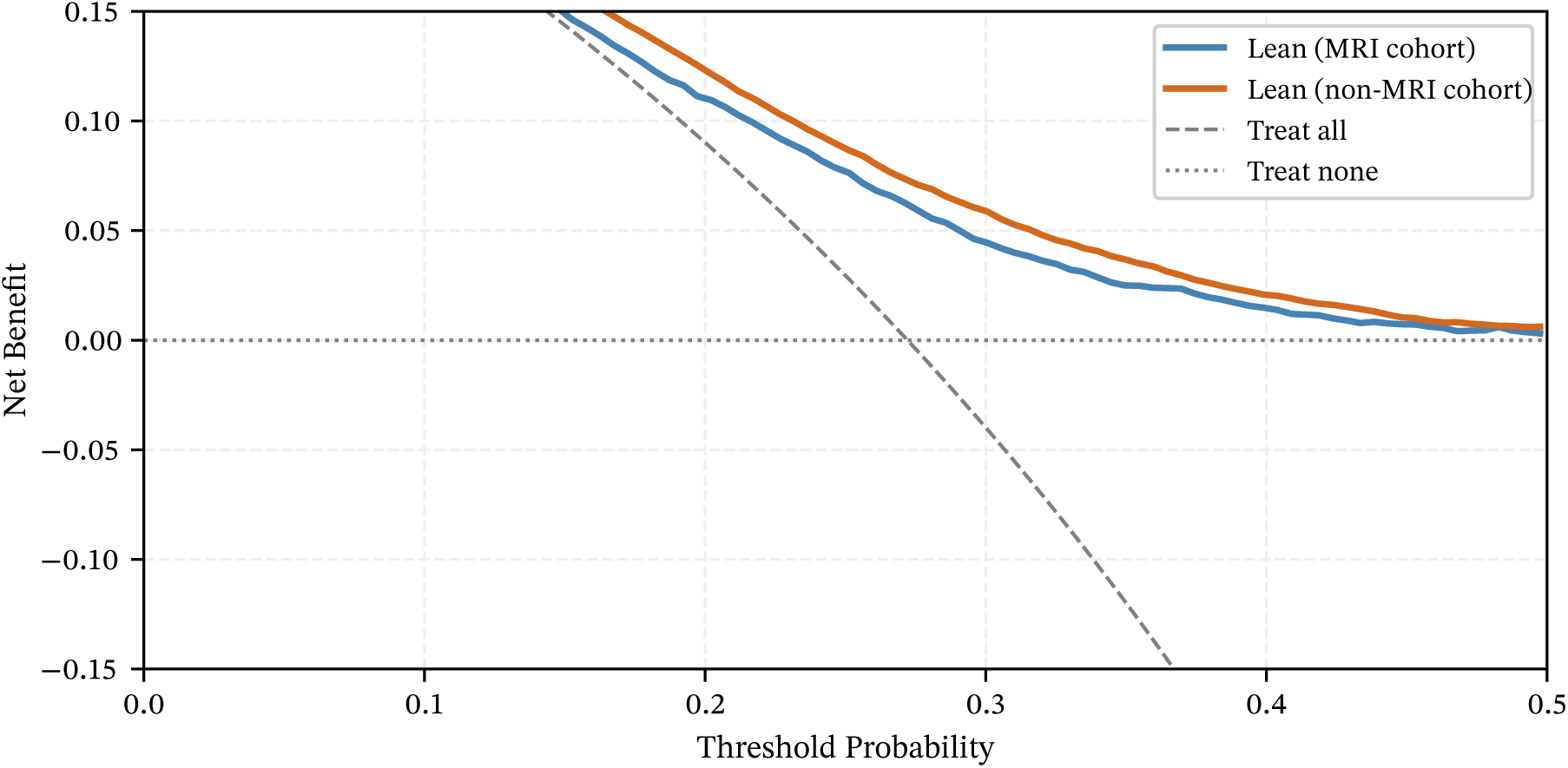
Decision-curve analysis for the Lean Universal Classifier in both NAKO cohorts. Net benefit across threshold probabilities for the four-modality Lean classifier (demographics, SES, medical history, baseline mental health) trained and evaluated separately on the MRI cohort (ROC-AUC 0.658) and the non-MRI cohort (ROC-AUC 0.664). The two curves track each other across the operationally relevant threshold range, so the four modalities support similar clinical utility in both cohorts. Because the two models were trained separately, the figure does not bear on transportability, which the no-refit transfer in Section 4.4 assesses.

## Does the acute infection modify the mental-health association?

The second-hit framing says a pre-infection vulnerability needs an infection to become PCC. We therefore tested whether a *more severe* infection strengthens the association, on three axes of the acute course, using the same composite baseline-mental-health exposure and the same age, sex and center adjustment as the other sensitivity analyses (Table S13). The axes are: hospitalization during the acute infection, on a ward or in intensive care, as reported at Corona-2; reinfection, meaning two or more reported infections against exactly one; and a first infection in or after January 2022, when Omicron became dominant in Germany. The last is a coarse contrast between pandemic periods and not a variant assignment: infection dates are month-precision, and a January-2022 infection may well have been Delta.

Three features of the design bound what these models can say. First, the acute course is recorded at Corona-2, that is *after*the exposure, so it is never a predictor in the classifier; these are effect-modification models on an established association, and adjusting for the acute course would mean conditioning on a post-exposure variable, the same objection the Discussion raises against adjusting for the Corona-2 symptom state. Second, hospitalization is rare: 66 participants in the MRI analytic sample, which yields an interval too wide to read (OR 1.17, 0.35–3.87). The pooled clean-controls cohort (*N* = 49,865, 14,045 PCC cases) is therefore the primary sample for this analysis. Third, hospitalization is itself a consequence of the infection and therefore downstream of the exposure, so stratifying on it inherits a weaker form of the same objection; the strata are read as descriptive contrasts, not as counterfactual comparisons.

Neither repeated infection (OR 0.98, 0.83–1.16) nor the pandemic era (1.11, 0.97–1.28) shows evidence of modification. The era result answers a question of its own: baseline mental health predicts PCC about equally in infections first reported in the Omicron period (OR 2.29) and before it (2.05), so the association is not an artifact of the early pandemic, even though the later era carries a markedly lower PCC prevalence (26.8% versus 33.6%).

Hospitalization does modify it, and in the direction opposite to a dose-response reading of the second hit. Among participants never hospitalized, the baseline-mental-health odds ratio is 2.25 (2.12–2.38); among the hospitalized it is 1.02 (0.56–1.83), i.e. indistinguishable from no association, with an interaction OR of 0.46 (0.26–0.81, *p* = 0.007).

The first alternative reading of that result is saturation rather than effect modification: PCC follows 57.9% of hospitalized infections against 27.9% otherwise, and where an outcome is already common an odds ratio has less room to move. That reading makes a specific prediction, namely a multiplicative interaction with no counterpart on the risk-difference scale, and it is not what the data show: the interaction is −0.171 on the additive scale as well (−0.305 to −0.036, *p* = 0.013). It also survives adjustment for the calendar timing of the infection, entered as months from December 2019 to the reported infection (OR 0.47, *p* = 0.009); because the survey window is fixed, that covariate stands in for how long participants had to recover before being asked.

The reading we favor is that the two routes to PCC are partly distinct: where the acute infection is severe enough to require admission, the illness itself supplies the risk, and the pre-infection psychiatric vulnerability adds little on top of it. Where the acute course is mild, which is the overwhelming majority of this cohort, the vulnerability is what separates those who go on to persistent symptoms from those who do not. That does not contradict a pre-infection vulnerability reading, but it bounds where such a reading could apply: what this paper documents is a predictor of PCC after mild COVID-19. Hospitalization is self-reported here, as the outcome is, and 361 exposed participants is thin for a subgroup claim, so we report the finding as a bound on generalization rather than as an established mechanism.

**Table S13:** Modification of the baseline-mental-health association by the acute course. Interaction between composite baseline-mental-health positivity and three features of the acute infection, on the pooled clean-controls cohort of the Lean refit (Section 4.4; *N* = 49,865, 14,045 PCC cases). Hospitalization status is missing for 177 participants, so that row’s models are fitted on 49,688. All models adjust for age, sex and study center. The multiplicative column is the interaction odds ratio from a logistic model; the additive column is the interaction risk difference from an identity-link model with heteroskedasticity-consistent (HC1) robust standard errors, reported because an interaction is scale-dependent and because saturation of a common outcome would produce the former without the latter. Stratum-wise odds ratios and PCC prevalences are given so that the saturation reading can be checked directly.

| Acute-course axis | *n* exposed | Interaction OR | Interaction risk diff. | MH OR by stratum<sup>a</sup> |
| --- | --- | --- | --- | --- |
| Hospitalized | 361 | 0.46 (0.26–0.81) | −0.171 (−0.305 to −0.036) | 1.02 vs. 2.25 |
| Reinfected | 5,715 | 0.98 (0.83–1.16) | +0.009 (−0.032 to +0.050) | 2.16 vs. 2.23 |
| Omicron era | 39,921 | 1.11 (0.97–1.28) | +0.012 (−0.022 to +0.045) | 2.29 vs. 2.05 |
| *PCC prevalence by stratum (exposed vs. unexposed)* |  |  |  |  |
| Hospitalized |  | 57.9% vs. 27.9% |  |  |
| Reinfected |  | 34.9% vs. 27.3% |  |  |
| Omicron era |  | 26.8% vs. 33.6% |  |  |

<sup>a</sup> Baseline-mental-health odds ratio fitted within the exposed and the unexposed stratum separately, same adjustment set. PCC cases per stratum (exposed vs. unexposed): 209 vs. 13,765 hospitalized, 1,992 vs. 12,053 reinfected, 10,701 vs. 3,344 Omicron era. The corresponding fits in the MRI-only cohort do not converge for the hospitalized and reinfected strata, which is a further consequence of how thin those strata are there.
PCC = Post-COVID Condition; MH = Mental Health; OR = odds ratio; HC1 = heteroskedasticity-consistent standard errors.

## Who the analytic sample lost

Of the 19,242 participants in the neuroimaging subsample, 10,778 (56.0%) do not reach the clean-controls analytic sample: 7,917 report no SARS-CoV-2 infection, 2,365 are infected but have no observable symptom outcome, and 496 are sub-threshold symptomatic and fall outside both arms by design. Because that is a majority of the imaged cohort, Table S14 compares the two groups on the baseline variables available for both. This is a different comparison from the MRI participation gradient of Table S3, which contrasts imaged with unimaged participants.

Only age separates them to any material degree: those excluded are on average four years older, which follows from the dominant exclusion reason, since reporting an infection is itself age-patterned. Sex is identical between the groups, and education and the two baseline mental-health scales differ by standardized mean differences below 0.1. The variables that carry this paper’s principal finding are therefore not the ones on which the analytic sample is selected.

**Table S14:** Participants entering the analytic sample against those excluded. Within the neuroimaging subsample (*N* = 19,242), baseline characteristics of the 8,464 participants entering the clean-controls analytic sample and the 10,778 excluded from it. Continuous variables are mean (SD); SMD is Cohen’s *d* for continuous and the standardized proportion difference for binary variables. *p*-values are from Welch’s *t*-test and the *χ*^2^ test; at these sample sizes the SMD is the informative summary and small *p*-values are expected for negligible differences.

| Variable | Included ($n = 8,464$) | Excluded ($n = 10,778$) | SMD | $p$ |
| --- | --- | --- | --- | --- |
| Age, years | 45.2 (11.7) | 49.3 (11.9) | -0.35 | <0.001 |
| Education (ISCED) | 4.5 (1.0) | 4.4 (1.0) | 0.07 | <0.001 |
| PHQ-9 baseline (0-27) | 3.5 (3.3) | 3.7 (3.6) | -0.05 | <0.001 |
| GAD-7 baseline (0-21) | 2.9 (2.8) | 3.0 (3.1) | -0.04 | 0.007 |
| Female sex | 3,700 (43.7%) | 4,728 (43.9%) | 0.00 | 0.844 |

SMD = standardized mean difference; SD = standard deviation; PHQ-9 = Patient Health Questionnaire-9; GAD-7 = Generalized Anxiety Disorder 7-item scale.

## Does baseline mental health predict measured olfaction, or only reported olfaction?

Loss of smell is the one PCC item this cohort measures twice: once by self-report in the Corona-2 survey, and once with an instrument, the 12-item Sniffin’ Sticks identification screening administered at the Level 2 follow-up examination. That makes the chemosensory domain the only place in this study where a self-reported outcome can be held against an instrument reading of the same construct (Section 3.3, item (vi)).

The contrast is paired: both outcomes are fitted on the same participants, with the same composite baseline-mental-health exposure and the same age, sex and center adjustment as the other sensitivity analyses, so the two odds ratios differ in what is measured and in nothing else. Timing decides what each can mean. For participants screened before their reported infection, the instrument cannot contain post-COVID-19 anosmia and measures the olfactory function they brought into the pandemic; if depressed participants had poorer olfaction all along, this is where it would show. For those screened afterwards, the instrument could register post-infection loss, but that stratum is small and is a convergence check rather than a result of its own.

Three limits bound the panel. The reconstructed examination year resolves to the calendar year only, so participants screened in their year of infection cannot be ordered against it and enter neither stratum. The reconstruction is available for part of the cohort only, so these models run on a subset of the infected clean-controls sample rather than all of it. And the objective classification is NAKO’s own, computed from the identification score; screenings NAKO declines to classify are treated as missing rather than as failed tests.

The pattern in Table S15 is mixed. Baseline mental health is unrelated to pre-infection measured hyposmia while predicting the later self-report in the same participants, and it is likewise unrelated to measured hyposmia among the never-infected; that is the reporting-style signature. Against it, the measured score predicts the self-report about as strongly as the PHQ-9 does, so the self-report is not detached from olfactory function, and in the post-infection stratum baseline mental health predicts the measured and the self-reported outcome at similar magnitude, which is what convergent olfactory vulnerability would look like. The post-infection interval starts at unity on 61 events, so that half of the pattern is suggestive rather than established. The Discussion therefore treats the chemosensory item as a further self-report outcome rather than a reporting-style-resistant one.

**Table S15:** Baseline mental health against measured and self-reported olfaction. Composite baseline-mental-health positivity as the exposure, adjusted for age, sex and study center, on infected clean-controls participants with a dated olfactory screening. The objective outcome is NAKO’s olfactory classification (hyposmia or anosmia against normosmia) from the 12-item Sniffin’ Sticks identification screening; the self-reported outcome is the Corona-2 loss-of-smell item. Within each stratum both outcomes are fitted on identical participants, so the two rows differ only in what is measured. Participants reporting a cold in the six weeks before the test are excluded. The ratio row is a percentile interval from 1,000 participant bootstrap resamples, of which 910 identified both models. The reporter-bias rows come from a linear-probability model regressing the self-report on the instrument reading and the baseline PHQ-9 together, reported as percentage points of self-report across the full range of each predictor so that the two are comparable.

| Outcome | *n* | Baseline-MH odds ratio [95% CI] |
| --- | --- | --- |
| *Screened before the reported infection* |  |  |
| Measured hyposmia | 7,227 | 0.89 [0.66, 1.19] |
| Self-reported loss of smell | 7,227 | 1.92 [1.22, 3.03] |
| Ratio, self-report / measured<sup>a</sup> | 7,227 | 2.12 [1.21, 3.57] |
| *Screened after the reported infection<sup>c</sup>* |  |  |
| Measured hyposmia | 506 | 2.12 [1.00, 4.52] |
| Self-reported loss of smell | 506 | 2.18 [0.83, 5.70] |
| *Reference: never infected* |  |  |
| Measured hyposmia | 22,969 | 1.06 [0.95, 1.19] |
| *What the self-report follows (linear-probability model, *n* = 6,990)<sup>b</sup>* |  |  |
| Measured identification score |  | −7.84 percentage points |
| Baseline PHQ-9 |  | +9.53 percentage points |

<sup>a</sup> Ratio of the self-report odds ratio to the measured-hyposmia odds ratio on the same participants; an interval excluding unity means the exposure predicts the two outcomes differently.
<sup>b</sup> Change in the probability of self-reporting loss of smell across the full range of each predictor (identification score 0–12; PHQ-9 sum 0–27), adjusted for age, sex and center. Both predictors enter the same model.
<sup>c</sup> This stratum rests on 61 measured-hyposmia and 34 self-reported events, which is why both intervals are wide and the measured-hyposmia interval starts at unity.
PCC = Post-COVID Condition; MH = Mental Health; CI = Confidence Interval; PHQ-9 = Patient Health Questionnaire-9.

## Footnotes

1 NAKO Corona-2 variable d_co2_k0: value 1 = persistent post-infection symptoms reported, value 2 = none reported; values 7775 (item not administered by the questionnaire routing) and 8888 (no answer) leave the outcome unobserved.

## Notes

### Competing Interest Statement

Christopher L. Schlett reports speaker fees from Siemens Healthineers, Bayer Healthcare and MSD, and a research grant from Siemens Healthineers. Fabian Bamberg reports consultancy and speaker fees and an unrestricted research grant from Bayer Healthcare, and an unrestricted research grant from Siemens Healthineers. Neither relationship is connected to the present analysis, which uses previously acquired NAKO imaging data and involves no industry funding, input or review. All other authors declare no competing interests.

### Author Declarations

The German National Cohort (NAKO Health Study) received ethical approval from the Ethics Committee of the Bavarian Medical Association (Bayerische Landesaerztekammer, Munich, Germany; protocol code 13023) as the lead vote, followed by the ethics committees responsible for each of the 18 NAKO study centres. The present work is a secondary analysis of pseudonymised NAKO data under the approved data use application NAKO-882 and required no additional participant contact. All participants gave written informed consent covering the baseline examination, the follow-up questionnaires and the scientific use of their pseudonymised data.

### Summary of Updates

Version 2 corrects data errors, reruns all analyses and adds one analysis. The main findings are unchanged: baseline mental health is the strongest pre-infection predictor, and brain MRI volumetry carried no predictive signal (ROC-AUC 0.665, previously 0.664). 1. Corrected variables. A check against the NAKO data dictionaries found three errors: the variable reported as hypertension was atopic eczema (physician-diagnosed hypertension now enters the model and Table 1), equivalized household income had been divided by the needs weight twice, and the cognitive score labelled number series was the Purdue pegboard. Discrimination moved in the third decimal at most. 2. Pipeline. A stability-selection stage retained every feature in every orthogonalized analysis and was removed. Its removal changed floating-point details that the meta-learner carries into calibration and operating points (calibration slope 1.016 to 1.030; sensitivity and specificity at the F1-optimal threshold 76.8 and 47.1 to 74.1 and 50.3 percent). 3. New analysis. Removing all six MRI atlases together and retraining the meta-learner leaves discrimination unchanged (ROC-AUC difference –0.002, 95% CI –0.004 to 0.000; Supplementary Table S7). The equivalence margin of 0.03 was fixed before this analysis but after the per-atlas MRI null of version 1 was known. 4. Further corrections. – Outcome: the PCS score rests on the 21 symptom items NAKO collected; version 1 also listed items that were not collected (such as brain fog and tinnitus). Supplementary Section S1 assigns each item to its PCS complex. – Transfer to the non-MRI cohort: the four-modality model meets all three equivalence criteria, the calibration intercept narrowly (upper limit 0.044; version 1: 0.0502, not met). The criterion is unchanged; the revised pipeline and corrected data moved the estimate. The transfer shows transportability within NAKO, not external validity. – The four modalities of the transferred model were chosen by availability; the rule of at least 0.02 PR-AUC over demographics no longer separates them. – Per-modality masking (Supplementary Table S6) is evaluated on held-out folds; ablation refers only to retraining without a modality. – The MRI null rests on a single neuroimaging cohort and does not show that COVID-19-associated brain changes are acute-onset. – E-value: 2.66 for the point estimate, 2.42 for the lower confidence limit. – Code: github.com/cl445/pcc-prediction-nako, release v1.2.0; version 1 cited a private repository. 5. Numbering. Supplementary Tables S7 to S14 are now S8 to S15; Sections S3 to S5 are now S2 to S4.

